# Functionally informed annotation influences pathway-specific polygenic risk and disease inference in Alzheimer’s disease

**DOI:** 10.64898/2026.05.25.26353905

**Authors:** Katrina Bazemore, Taha Iqbal, Amanda B Kuzma, Struan F. A. Grant, Gerard D. Schellenberg, Li-San Wang, Alessandra Chesi, Jin Jin, Adam C. Naj

**Author notes:** Corresponding author: Katrina Bazemore Corresponding author’s.

## Abstract

Pathway-specific polygenic risk scores (pathway-PRS) measure aggregate risk across single nucleotide variants (SNPs) annotated to pathway genes. In most applications, SNP-to-gene annotation is based on SNP proximity to gene boundaries. This approach is ill-suited for incorporating non-coding SNPs, which can regulate gene expression over long distances and represent a large proportion of risk variants in complex diseases, such as Alzheimer’s disease (AD). AD therefore provides a useful setting for evaluating whether functionally informed SNP-to-gene annotation improves pathway-PRS construction. Here, we compare AD pathway-PRS performance across annotation strategies that integrate varying levels of functional genomic data, including adult brain chromatin interaction and expression quantitative trait loci (eQTL) data. In the UK Biobank (n=328,526), including AD cases defined by ICD-9/10 codes (n=3,043) and family history of AD/dementia (n=38,589), the strategy integrating chromatin interaction and eQTL data consistently improves pathway-PRS performance. We replicate this finding in independent Alzheimer’s Disease Genetics Consortium data (n=3,370). We further observe that pathway-PRS associations with AD vary by annotation strategy and that integrative annotation increases power to detect sex-dependent and age-at-onset associations. Together, these findings support the use of functionally informed SNP-to-gene annotation for pathway-PRS construction and highlight the importance of applying multiple annotation strategies for robust inference.

## Introduction

Alzheimer’s disease (AD) is a debilitating and complex disease that affects an estimated 6 million people in the US^1^. Efforts to understand the genetic basis of AD have been complicated by its clinical and genetic heterogeneity^2^. Pathway-specific polygenic risk scores (pathway-PRS) have recently been applied to AD to investigate the contribution of biological pathways to this heterogeneity^3,4^. In contrast to global PRS, which estimate aggregate risk genome-wide^5,6^, pathway-PRS estimate aggregate genetic risk for a disease within a biological pathway, allowing for disease subtyping, insight into pathway roles in endophenotypes or pathogenesis, and more precise examinations of gene-risk factor interactions^7^. Pathway-PRS have been used in the AD context to identify pathway-specific associations with cognitive-decline^3,8^ or AD risk^9^ and resilience^4^, predict changes in fluid biomarkers^10^, identify sex-dependent pathway-trait associations^11^, explore mediation of disease risk by a pathway-relevant biomarker^12^, and link biological pathways with endophenotypes^13,14^. Given the increasing application of pathway-PRS in efforts to understand genetically driven AD pathogenesis, it is essential to re-examine the methods used to construct them.

A key aspect of pathway-PRS construction is the annotation of SNPs to pathway genes. Pathway-PRS are typically built by (1) identifying genes in the pathway of interest, (2) defining a set of single-nucleotide polymorphisms (SNPs) relevant to the function and regulation of pathway genes, and (3) training a weighted linear combination of these SNPs. SNP-to-gene annotation, the second step in this process, often receives little attention but has important implications for pathway-PRS interpretability and predictive performance. It is well established that complex trait risk loci often reside in non-coding regions and colocalize with elements that regulate gene expression at a distance^15^. Previously, non-coding risk loci identified in genome-wide association studies (GWAS) were widely assumed to regulate the expression of the nearest gene^16^. This assumption underpins the current, position-based, annotation strategies used in many pathway-PRS analyses, which typically assign SNPs to pathway genes if they fall within 10-50 kilobase (kb) windows around gene boundaries^7–10,12,17^. However, multiple investigations indicate that a substantial proportion of effector genes lie more than 50kb from their corresponding GWAS loci^18–20^. Positional annotation therefore excludes elements that regulate pathway gene expression and instead includes irrelevant regions, potentially diluting pathway associations or, worse still, attributing association signals to the wrong biological mechanism.

While some studies aim to address this issue by using wider windows for positional annotation^21^, adding SNPs associated with pathway gene expression (eQTLs)^4,13,22^, or performing manual annotation based on literature reviews^3,4,13^, few studies have incorporated genome-wide, integrative, and systematic methods to annotate distal regulatory elements to pathway genes. Fortunately, publicly available data from projects such as GTEx^23^ and PsychENCODE^20^ have enabled comprehensive tissue-specific SNP-to-gene annotation, as implemented in integrative pathway enrichment analysis tools such as *H-MAGMA*^24,25^. Comparison of such SNP-to-gene annotation strategies has not yet been performed in the pathway-PRS context, though there is great potential for more integrative strategies to improve pathway-PRS predictive performance.

Thus, in this study we sought to assess the utility of various approaches for constructing AD pathway-PRS, with the aim of demonstrating the potential of systematic SNP-to-gene annotation integrating functional data. First, we compared performance of pathway-PRS constructed using three different SNP-to-gene annotation strategies. Next, we tested a method to prioritize inclusion of SNPs in open chromatin regions and examined the potential of alternately defined enhancer-gene pairs^19^ to improve the accessibility of integrative annotation. Finally, we repeated our performance comparisons in an independent dataset with rigorous ascertainment of AD status, replicating our key findings, demonstrating increased power in association testing with endophenotypes and in stratified analyses, and showing how SNP-to-gene annotation can influence conclusions drawn from pathway-PRS analyses.

## Results

### Study Participants

Our study participants were drawn from two independent datasets, the UK Biobank (UKB)^26^ and the Alzheimer’s Disease Genetics Consortium (ADGC)^27–29^. We first examined pathway-PRS predictive performance by annotation strategy in the UKB. Our proxy analysis included participants with true and ‘proxy AD’ (see Methods) as cases and those aged 50 years or older with no other dementia diagnoses as controls (n=328,526, median age 72.8 years, 54.77% female, 12.8% with ≥1 parent affected; **Table 1; Supplementary Fig. 1**). We also included a true case analysis (n=259,418, median age 74.30 years, 53.66% female, 14.48% with ≥1 parent affected; **Supplementary Fig. 2**) and an analysis based on the methods of Wu et al. 2024^30^ and Marioni et al. 2018^31^ (n=202,408, median age 72.90 years, 54.76% female, 16.44% with ≥1 parent affected; **Supplementary Fig. 3**; see Methods). Second, we calculated global and pathway-PRS trained on the UKB in a set of 3,370 European ancestry AD cases and controls from the ADGC (median age 74.00 years, 58.10% female; **Table 1**).

**Table 1.**
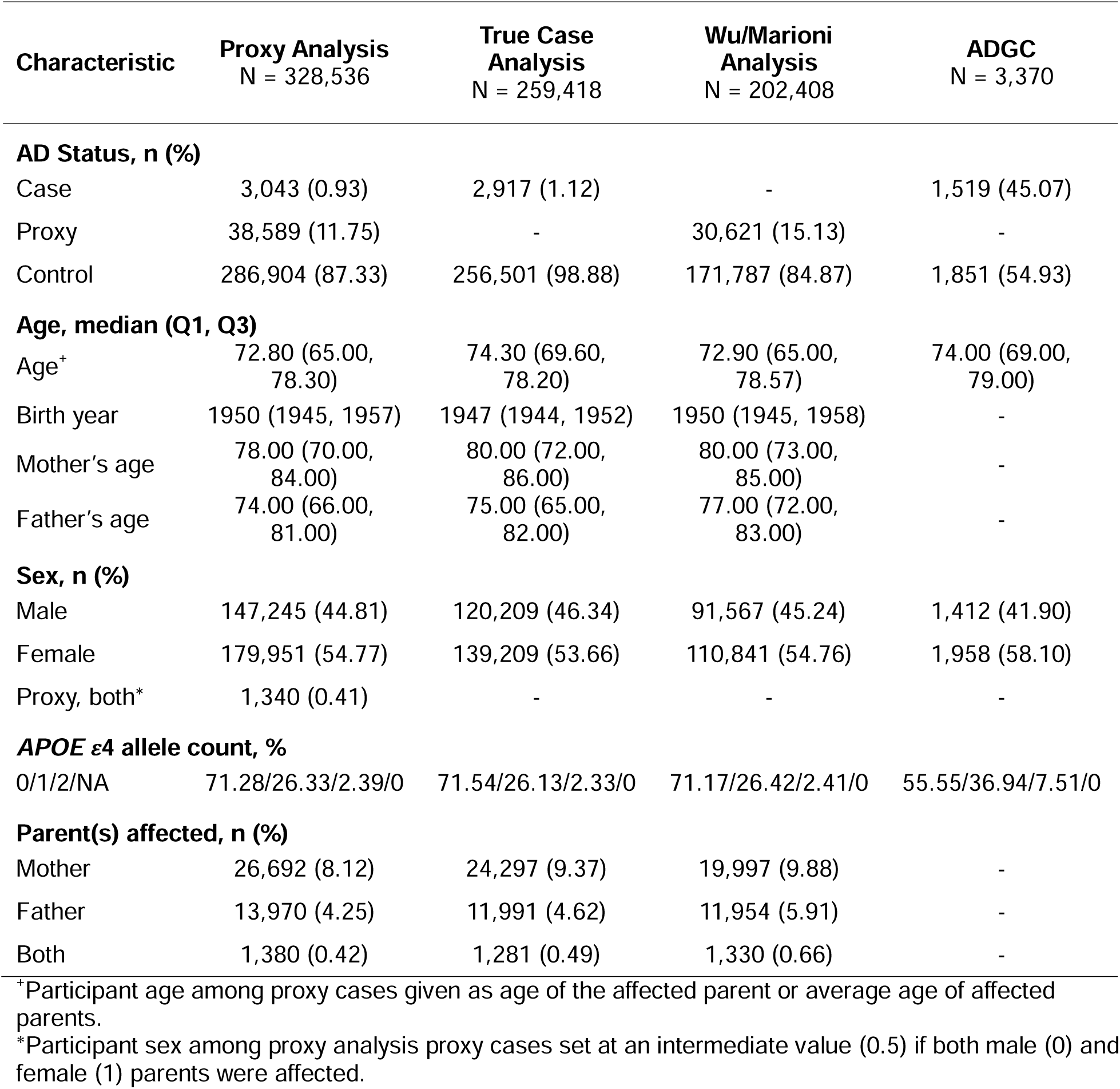
Participant Characteristics.

| Characteristic | Proxy Analysis<br>N = 328,536 | True Case<br>Analysis<br>N = 259,418 | Wu/Marioni<br>Analysis<br>N = 202,408 | ADGC<br>N = 3,370 |
| --- | --- | --- | --- | --- |
| **AD Status, n (%)** |  |  |  |  |
| Case | 3,043 (0.93) | 2,917 (1.12) | - | 1,519 (45.07) |
| Proxy | 38,589 (11.75) | - | 30,621 (15.13) | - |
| Control | 286,904 (87.33) | 256,501 (98.88) | 171,787 (84.87) | 1,851 (54.93) |
| **Age, median (Q1, Q3)** |  |  |  |  |
| Age<sup>+</sup> | 72.80 (65.00, 78.30) | 74.30 (69.60, 78.20) | 72.90 (65.00, 78.57) | 74.00 (69.00, 79.00) |
| Birth year | 1950 (1945, 1957) | 1947 (1944, 1952) | 1950 (1945, 1958) | - |
| Mother's age | 78.00 (70.00, 84.00) | 80.00 (72.00, 86.00) | 80.00 (73.00, 85.00) | - |
| Father's age | 74.00 (66.00, 81.00) | 75.00 (65.00, 82.00) | 77.00 (72.00, 83.00) | - |
| **Sex, n (%)** |  |  |  |  |
| Male | 147,245 (44.81) | 120,209 (46.34) | 91,567 (45.24) | 1,412 (41.90) |
| Female | 179,951 (54.77) | 139,209 (53.66) | 110,841 (54.76) | 1,958 (58.10) |
| Proxy, both* | 1,340 (0.41) | - | - | - |
| **APOE ε4 allele count, %** |  |  |  |  |
| 0/1/2/NA | 71.28/26.33/2.39/0 | 71.54/26.13/2.33/0 | 71.17/26.42/2.41/0 | 55.55/36.94/7.51/0 |
| **Parent(s) affected, n (%)** |  |  |  |  |
| Mother | 26,692 (8.12) | 24,297 (9.37) | 19,997 (9.88) | - |
| Father | 13,970 (4.25) | 11,991 (4.62) | 11,954 (5.91) | - |
| Both | 1,380 (0.42) | 1,281 (0.49) | 1,330 (0.66) | - |

<sup>+</sup>Participant age among proxy cases given as age of the affected parent or average age of affected parents.
\*Participant sex among proxy analysis proxy cases set at an intermediate value (0.5) if both male (0) and female (1) parents were affected.

### Annotation of SNPs to Pathway Clusters

To examine the impact of SNP-to-gene annotation on pathway-PRS predictive performance, we constructed AD pathway-PRS under three annotation strategies. We selected a set of AD pathways using *MAGMA*^32^ pathway enrichment analysis of the 2019 International Genomics of Alzheimer’s Project GWAS (IGAP GWAS)^29^, excluding the *APOE* region^17^. We identified 37 Gene Ontology (GO) pathways which collapsed into 20 clusters based on shared gene content (see Methods), with 11 pathways forming singleton clusters (**Table 2**). The number of genes in each cluster ranged from 11 to 757 (**Fig. 1a**).

**Figure 1.**
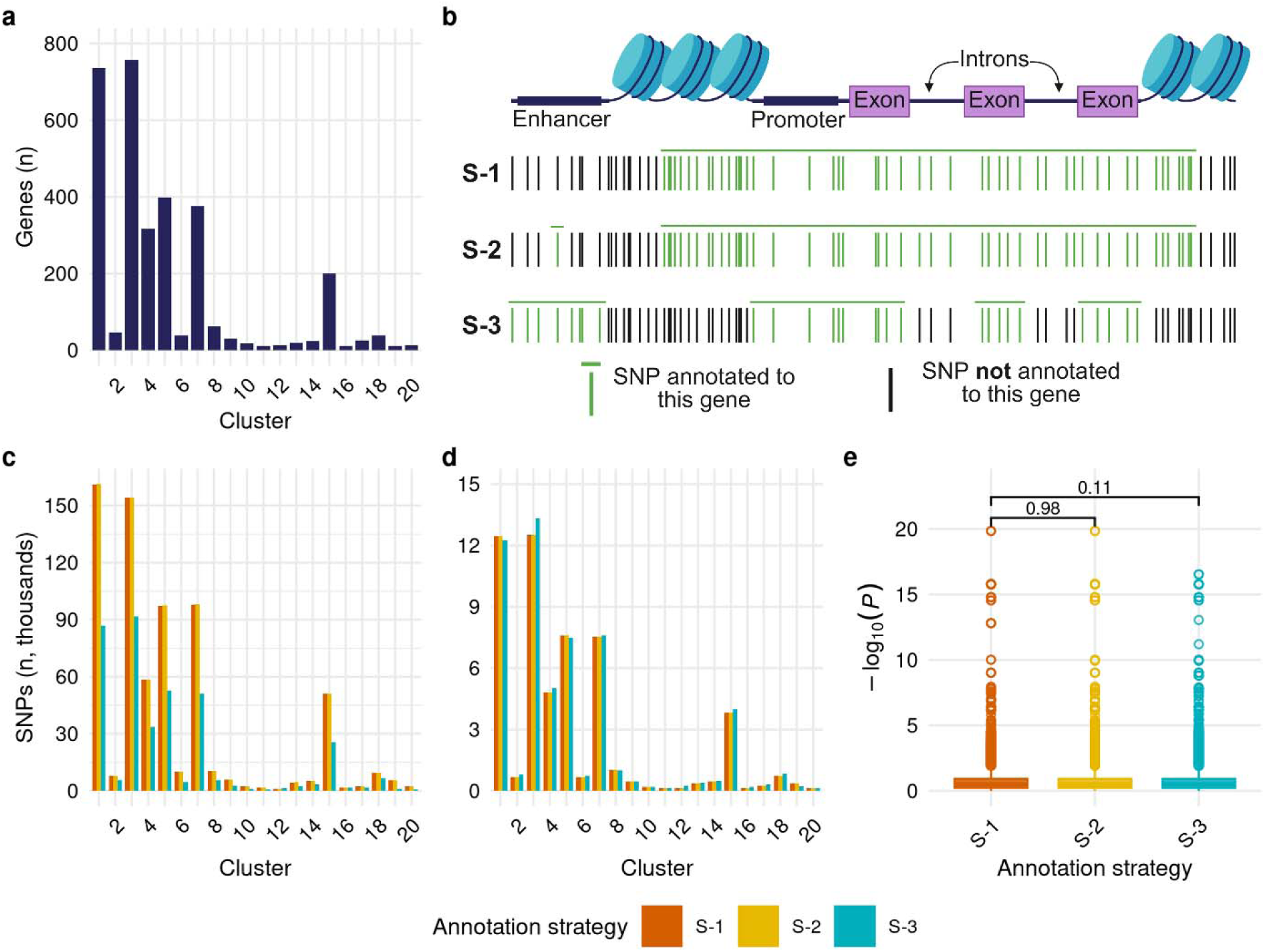
Characterization of SNP-to-gene annotation. **(a)** Number of unique protein-coding genes within each cluster, derived from GO pathway annotations. **(b)** Representation of SNPs captured by each annotation strategy for a given gene^+^. **(c)** Number of unique SNPs annotated to each cluster under each annotation strategy in thousands. **(d)** Number of unique linkage disequilibrium (LD)-independent SNPs (*r^2^* = 0.1) annotated to each cluster under each annotation strategy in thousands. **(e)** *P*-value distributions of LD-independent annotated variants pooled over all clusters by annotation strategy. Wilcoxon rank sum test \*\*\**p* < 0.001, \*\**p* < 0.01, \**p* < 0.05. ^+^Created in BioRender. Bazemore, K. (2026) https://BioRender.com/giotb5e.

**Table 2.**
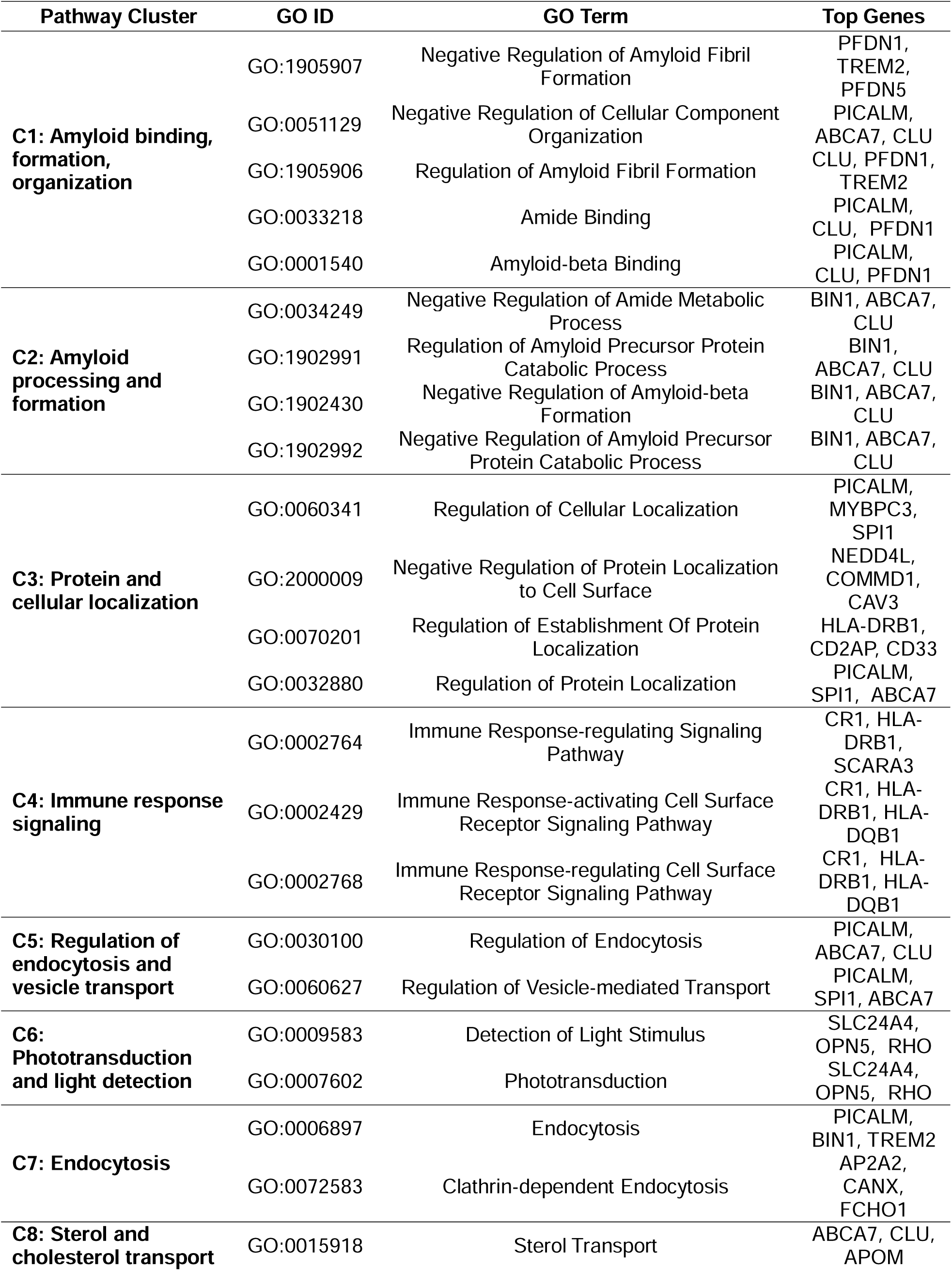
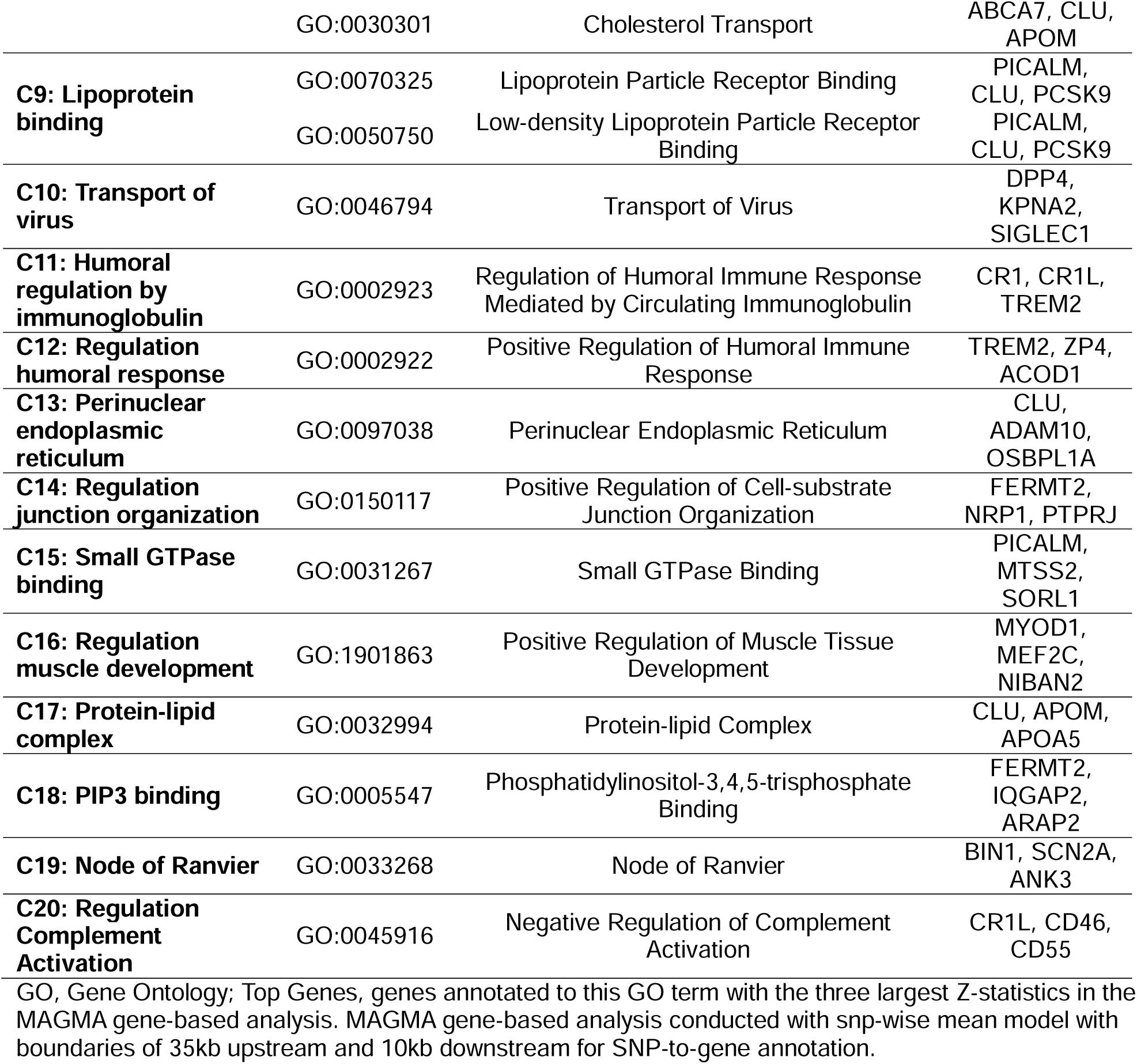
Alzheimer’s disease pathway enrichment and pathway clusters.

| Pathway Cluster | GO ID | GO Term | Top Genes |
| --- | --- | --- | --- |
| **C1: Amyloid binding, formation, organization** | GO:1905907 | Negative Regulation of Amyloid Fibril Formation | PFDN1, TREM2, PFDN5 |
|  | GO:0051129 | Negative Regulation of Cellular Component Organization | PICALM, ABCA7, CLU |
|  | GO:1905906 | Regulation of Amyloid Fibril Formation | CLU, PFDN1, TREM2 |
|  | GO:0033218 | Amide Binding | PICALM, CLU, PFDN1 |
|  | GO:0001540 | Amyloid-beta Binding | PICALM, CLU, PFDN1 |
| **C2: Amyloid processing and formation** | GO:0034249 | Negative Regulation of Amide Metabolic Process | BIN1, ABCA7, CLU |
|  | GO:1902991 | Regulation of Amyloid Precursor Protein Catabolic Process | BIN1, ABCA7, CLU |
|  | GO:1902430 | Negative Regulation of Amyloid-beta Formation | BIN1, ABCA7, CLU |
|  | GO:1902992 | Negative Regulation of Amyloid Precursor Protein Catabolic Process | BIN1, ABCA7, CLU |
| **C3: Protein and cellular localization** | GO:0060341 | Regulation of Cellular Localization | PICALM, MYBPC3, SPI1 |
|  | GO:2000009 | Negative Regulation of Protein Localization to Cell Surface | NEDD4L, COMMD1, CAV3 |
|  | GO:0070201 | Regulation of Establishment Of Protein Localization | HLA-DRB1, CD2AP, CD33 |
|  | GO:0032880 | Regulation of Protein Localization | PICALM, SPI1, ABCA7 |
| **C4: Immune response signaling** | GO:0002764 | Immune Response-regulating Signaling Pathway | CR1, HLA-DRB1, SCARA3 |
|  | GO:0002429 | Immune Response-activating Cell Surface Receptor Signaling Pathway | CR1, HLA-DRB1, HLA-DQB1 |
|  | GO:0002768 | Immune Response-regulating Cell Surface Receptor Signaling Pathway | CR1, HLA-DRB1, HLA-DQB1 |
| **C5: Regulation of endocytosis and vesicle transport** | GO:0030100 | Regulation of Endocytosis | PICALM, ABCA7, CLU |
|  | GO:0060627 | Regulation of Vesicle-mediated Transport | PICALM, SPI1, ABCA7 |
| **C6: Phototransduction and light detection** | GO:0009583 | Detection of Light Stimulus | SLC24A4, OPN5, RHO |
|  | GO:0007602 | Phototransduction | SLC24A4, OPN5, RHO |
| **C7: Endocytosis** | GO:0006897 | Endocytosis | PICALM, BIN1, TREM2 |
|  | GO:0072583 | Clathrin-dependent Endocytosis | AP2A2, CANX, FCHO1 |
| **C8: Sterol and cholesterol transport** | GO:0015918 | Sterol Transport | ABCA7, CLU, APOM |
|  | GO:0030301 | Cholesterol Transport | ABCA7, CLU, APOM |
| **C9: Lipoprotein binding** | GO:0070325 | Lipoprotein Particle Receptor Binding | PICALM, CLU, PCSK9 |
|  | GO:0050750 | Low-density Lipoprotein Particle Receptor Binding | PICALM, CLU, PCSK9 |
| **C10: Transport of virus** | GO:0046794 | Transport of Virus | DPP4, KPNA2, SIGLEC1 |
| **C11: Humoral regulation by immunoglobulin** | GO:0002923 | Regulation of Humoral Immune Response Mediated by Circulating Immunoglobulin | CR1, CR1L, TREM2 |
| **C12: Regulation humoral response** | GO:0002922 | Positive Regulation of Humoral Immune Response | TREM2, ZP4, ACOD1 |
| **C13: Perinuclear endoplasmic reticulum** | GO:0097038 | Perinuclear Endoplasmic Reticulum | CLU, ADAM10, OSBPL1A |
| **C14: Regulation junction organization** | GO:0150117 | Positive Regulation of Cell-substrate Junction Organization | FERMT2, NRP1, PTPRJ |
| **C15: Small GTPase binding** | GO:0031267 | Small GTPase Binding | PICALM, MTSS2, SORL1 |
| **C16: Regulation muscle development** | GO:1901863 | Positive Regulation of Muscle Tissue Development | MYOD1, MEF2C, NIBAN2 |
| **C17: Protein-lipid complex** | GO:0032994 | Protein-lipid Complex | CLU, APOM, APOA5 |
| **C18: PIP3 binding** | GO:0005547 | Phosphatidylinositol-3,4,5-trisphosphate Binding | FERMT2, IQGAP2, ARAP2 |
| **C19: Node of Ranvier** | GO:0033268 | Node of Ranvier | BIN1, SCN2A, ANK3 |
| **C20: Regulation Complement Activation** | GO:0045916 | Negative Regulation of Complement Activation | CR1L, CD46, CD55 |

GO, Gene Ontology; Top Genes, genes annotated to this GO term with the three largest Z-statistics in the MAGMA gene-based analysis. MAGMA gene-based analysis conducted with snp-wise mean model with boundaries of 35kb upstream and 10kb downstream for SNP-to-gene annotation.

SNPs reported in the IGAP GWAS^29^ summary statistics were annotated to cluster genes. Prioritized IGAP GWAS SNPs were SNPs in linkage disequilibrium (LD) (*r^2^*≥0.6) with a lead SNP (*P*<10^-5^, LD-independent at *r^2^*=0.6). Annotation was performed as follows (**Fig. 1b**):

**Strategy 1 (S-1):** based on SNP position within 35kb upstream or 10kb downstream of gene boundaries^33^;

**Strategy 2 (S-2):** S-1 plus annotation of the prioritized subset of SNPs to putative target genes based on adult brain eQTL and high-throughput chromosome (Hi-C) conformation capture data in *FUMA GWAS*^34^;

**Strategy 3 (S-3):** based on SNP position within gene exon or promoter regions plus annotation of any IGAP GWAS SNPs to putative target genes based on adult brain eQTL and Hi-C data with the *H-MAGMA* pipeline^25,33,35,36^.

S-1 represents a typical strategy applied in pathway-PRS studies. S-2 extends the typical strategy by incorporating functional annotation for a set of prioritized SNPs, analogous to targeted manual annotation based on a literature review. S-3 is a data-driven strategy that systematically includes SNPs with evidence of relevance to gene expression or function.

There was little difference in the number of SNPs annotated to clusters under S-1 or S-2 (median difference=0.03% [IQR:0%, 0.28%]). Under S-3, there were fewer SNPs annotated to clusters than under S-1 (median difference=-46.03% [IQR:-35.03%, -50.98%]; **Fig. 1c**). Clumping was performed to obtain annotated SNPs representing LD-independent loci. The number of LD-independent SNPs was similar across annotation strategies, with a median difference between S-1 and S-3 of 5.23% (IQR:-1.23%, 8.07%), indicating that S-1/S-2 capture more SNPs within loci, but not more loci than S-3 (**Fig. 1d**). There was no significant differences in the distribution of GWAS *p-*values for pooled LD-independent SNPs by annotation strategy (**Fig. 1e**). For a single cluster, C4: *Immune response signaling*, we observed a significant difference in the *p-*value distribution of LD-independent SNPs across annotation strategies (median - log(*p*)_S-1/S-2_= 0.526, median -log(*p*)_S-3_=0.494, *p=*0.018; **Supplementary Fig. 4**) indicative of S-3 capturing SNPs with less evidence of association with AD than S-1.

### Integrative SNP-to-gene annotation improves pathway-PRS performance in the UKB

We trained and tested global and pathway-PRS in the UKB across 10 repeated 80:20 holdouts in proxy and true case analyses. In the 80% training set, we applied clumping and thresholding (C+T) in *PRSet*^7^ to select best-fit global and pathway-PRS (see Methods). The *p-*value threshold yielding the best-fit PRS tended to be higher under the S-3 annotation strategy than under S-1 or S-2, retaining SNPs with less evidence of association with AD (**Supplementary Fig. 5**). In the testing sets, AD status was regressed on covariates and best-fit PRS. Standardized mean differences of AD status, age, sex, *APOE* ε2/ε3/ε4 haplotype distribution, and number of affected parents across training and testing sets were <0.01 for each repeated holdout. The correlations between pathway-PRS remained stable across annotation strategies (**Supplementary Fig. 6**).

Notably, several pathway-PRS outperformed the global PRS in both proxy and true case analyses on the incremental *R*^2^ metric measuring the variance in AD status explained by the PRS (see Methods). When constructed with S-3 annotation, the cluster 2 (C2): *Amyloid processing and formation* pathway-PRS outperformed the global PRS in the proxy analysis (Figure 2a-b). In the true case analysis, three pathway-PRS outperformed the global PRS under all annotation strategies, five pathway-PRS outperformed the global PRS only under S-3 and one only under S-1/S-2 (**Fig. 2c-d**). Thus, while pathway-PRS represent a subset of risk loci, they provide a powerful and biologically interpretable method for measurement of genetic risk and are further enhanced by the integration of functional genomics.

**Figure 2.**
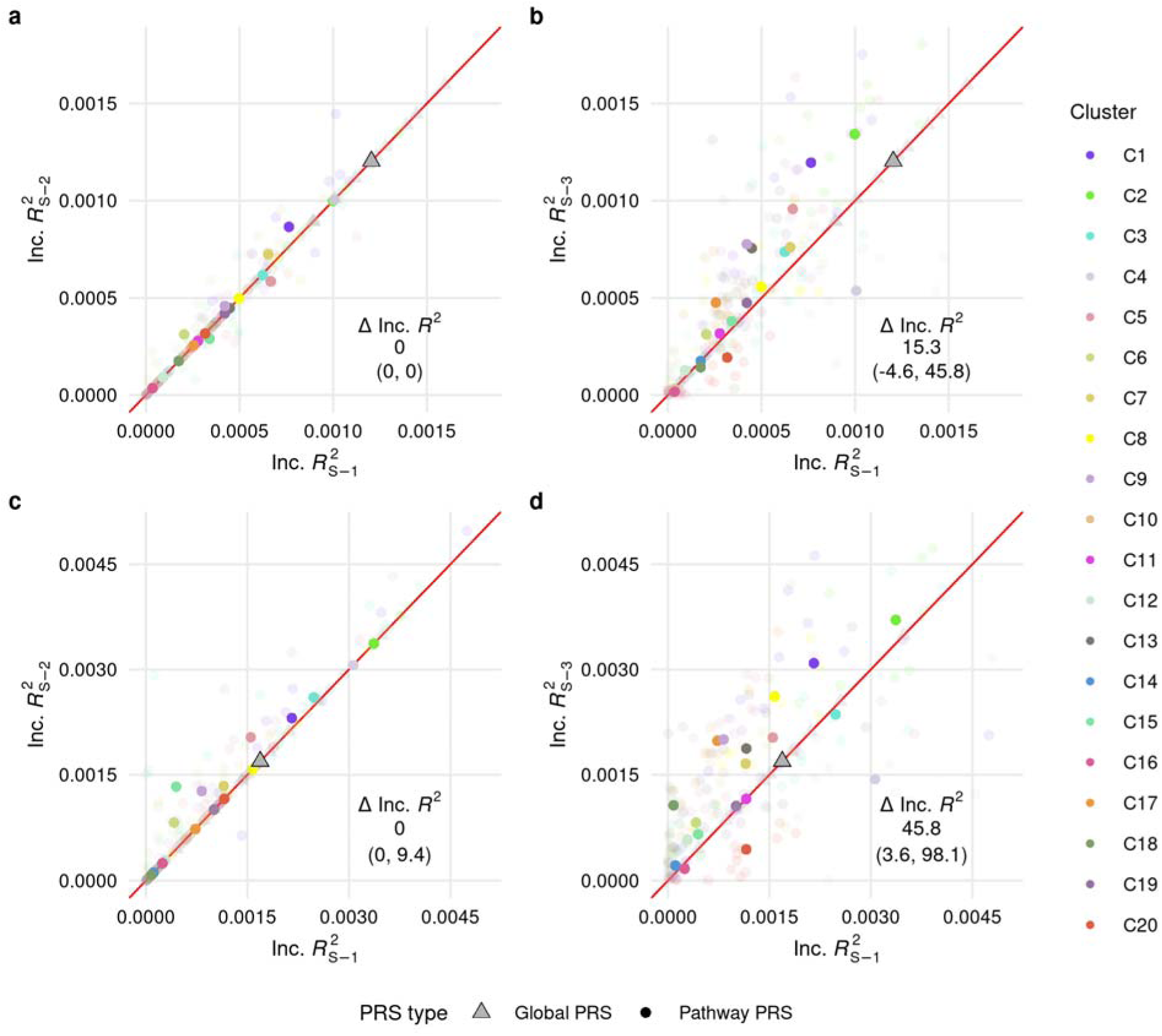
Global and pathway-PRS predictive performance. Performance in the UK Biobank testing sets under **(a&c)** S-1 vs S-2 annotation, **(b&d)** S-1 vs S-3 annotation. Top row shows performance in the proxy analysis; bottom row shows true case analysis. Inc.*R_X_*^2^, incremental *R*^2^ for each PRS as full model compared to covariate-only model; ΔInc.*R*^2^, percent change in Inc.*R*^2^ as median (Q1, Q3); transparent points, Inc.*R*^2^ estimates from individual holdouts; solid points, average Inc.*R*^2^ over ten repeated holdouts. One plot point with single holdout values was removed from (c&d) for C2 and C8 due to extreme values (C2 S-1: 0.0077, S-2: 0.0077, S-3: 0.0076; C8 S-1: 0.0033, S-3: 0.0071).

The predictive performance of pathway-PRS was highest when using integrative SNP-to-gene annotation. The S-2 annotation strategy yielded little difference in pathway-PRS predictive performance compared to S-1 (**Fig. 2a&c**), but performance was substantially elevated under S-3 (Figure 2b&d). Out of 20 clusters, 14 and 15 had increased pathway-PRS performance under S-3 in the proxy and true case analyses, respectively. Relative differences in performance were larger in the true case analysis (median:45.8% [IQR:3.6%, 98.1%]) than the proxy analysis (15.3% [IQR:-4.6%, 45.8%]) but tended to be less stable across repeated holdouts due to the smaller number of cases (**Fig. 2b&d**). Three pathway-PRS had reduced performance under S-3 in both analyses (C4, C10, C20). C4 and C20 had low absolute incremental *R^2^* values, indicating that they explain a small proportion of the AD variance relative to other pathway-PRS. The C1: *Amyloid binding, formation, organization* and C2: *Amyloid processing and formation* pathway-PRS explained the greatest variance in the AD outcome under S-3 and showed marked performance increases with S-3, highlighting the potential of S-3 to improve pathway-PRS with high trait relevance.

Improvement in incremental *R*^2^ translated to increased association strength and better risk stratification. The magnitude of pathway-PRS associations with AD risk were elevated under S-3 for 15 pathway-PRS in the proxy analysis and 16 in the true case analysis (**Fig. 3**). Pathway-PRS constructed with S-3 annotation additionally better distinguished risk in the extremes of the PRS distribution as shown by the greater prevalence of AD in the top decile and lower prevalence in the bottom decile of pathway-PRS constructed with S-3 than with S-1 (**Fig. 4**). Thus, the S-3 annotation strategy improves power for downstream analyses and enhances risk stratification.

**Figure 3.**
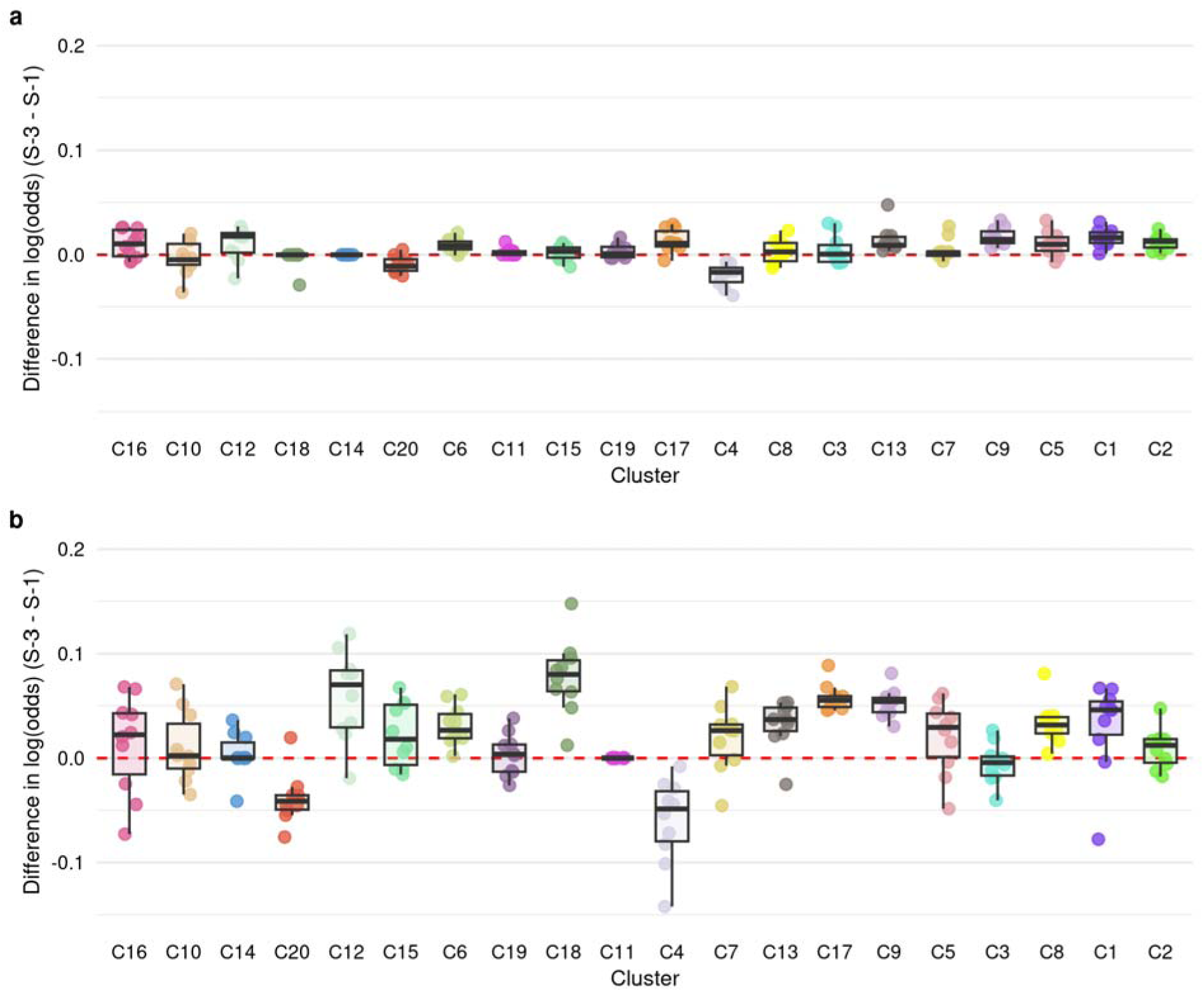
Change in magnitude of pathway-PRS associations with AD. Change in association magnitude between S-1 and S-3 annotation strategies across 10 repeated holdout testing sets in the **(a)** proxy analysis and **(b)** true case analysis adjusted for age, sex, number of *APOE ε*4 alleles, and top PCs. Clusters ordered by increasing pathway-PRS incremental R^2^ under S-3.

**Figure 4.**
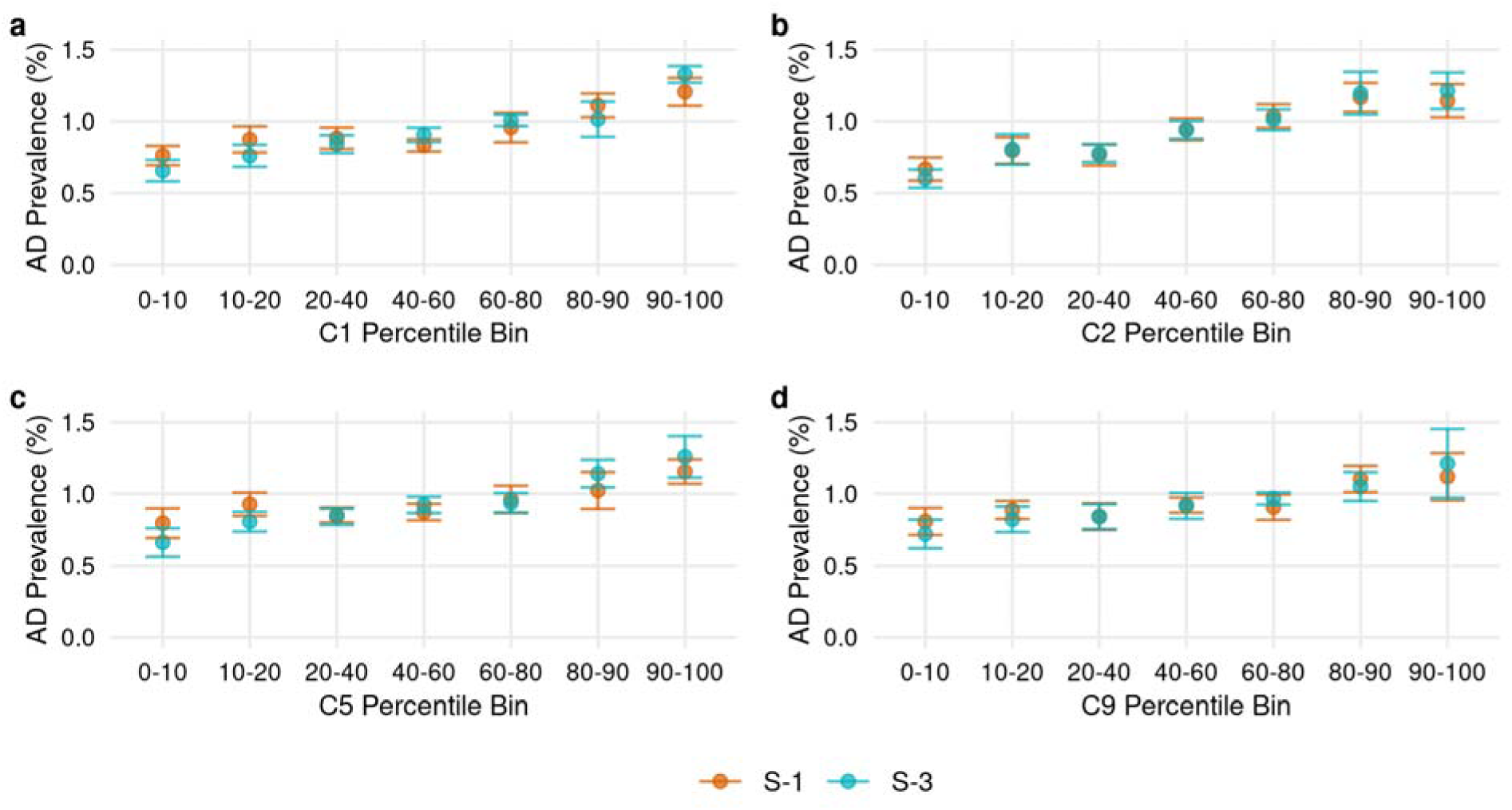
AD risk stratification across annotation strategies. Prevalence of true AD in bins of increasing pathway-PRS within UKB proxy analysis testing sets under annotation strategies S-1 and S-3 for clusters **(a)** C1: *Amyloid binding, formation, organization*, **(b)** C2: *Amyloid processing and formation*, **(c)** C5: *Regulation of endocytosis and vesicle transport*, **(d)** C9: *Lipoprotein binding.* Mean (points) and standard deviation (error bars) of prevalence across 10 repeated holdouts.

### Performance gains are robust to AD definition and prioritization of SNPs in open chromatin

We conducted sensitivity analyses to examine the consistency of our results under different AD outcome definitions, prioritization of active regions across all annotation strategies, and alternately defined enhancer-gene pairs. As proxy cases may introduce bias, we first conducted a sensitivity analysis excluding participants with missing parental AD/dementia status or whose parents were under 65 years of age (Wu/Marioni analysis; see Methods).

Results were consistent across AD outcome definitions (**Fig. 5a**). Pathway-PRS predictive performance was elevated under S-3, but not S-2 annotation across analyses (**Supplementary Fig. 7**). The median relative difference in pathway-PRS performance under S-3 in the sensitivity analysis was similar to the true case analysis (True case:45.8% [IQR:3.6%, 98.1%], Wu/Marioni:49.1% [IQR:4.0%, 72%]). While the magnitude of improvement varied, the pattern of pathway-PRS performance under S-3 generally remained consistent across analyses, with the exception of C18: *PIP3 binding*, which had increased predictive performance only in the true case analysis (**Fig. 3, Supplementary Fig. 8**).

**Figure 5.**
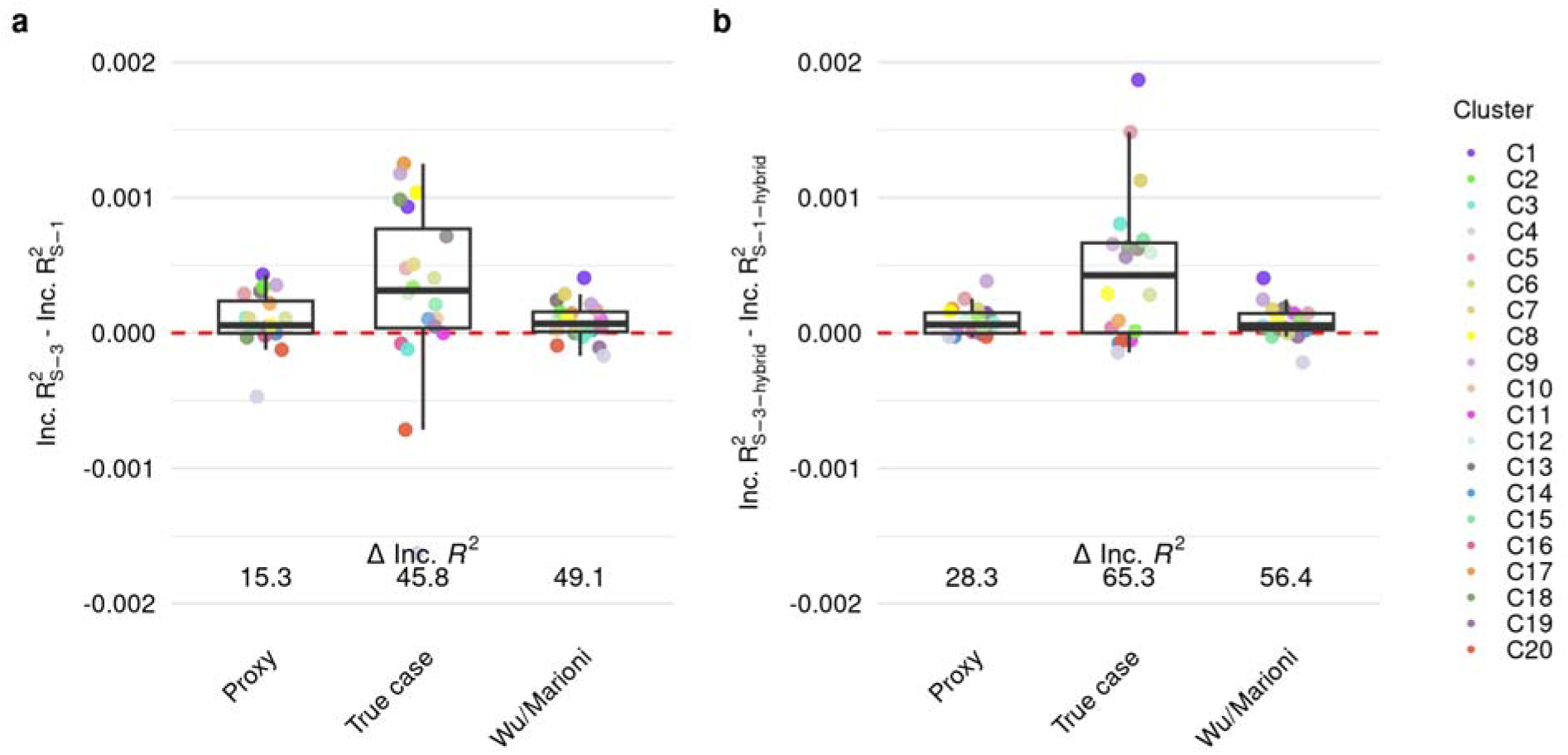
Pathway-PRS performance across outcome definitions. Difference in pathway-PRS predictive performance as averaged incremental *R*^2^ across repeated holdout testing sets between **(a)** standard C+T on S-1 and S-3 annotations, **(b)** hybrid C+T on S-1 and S-3 annotations. Training and testing conducted on (Proxy) proxy analysis dataset with adjustment for participant age*, sex*, number of APOE ε4 alleles, and top PCs, (True case) true case analysis dataset with adjustment for participant age, sex, number of APOE ε4 alleles, and top PCs, (Wu/Marioni) Wu/Marioni analysis dataset with adjustment for participant birth year, sex, maternal age, paternal age, genotyping batch, number of APOE ε4 alleles, and top PCs Inc.*R_X_*^2^, incremental *R*^2^ for each PRS as full model compared to covariate-only model, averaged over ten repeated holdouts; ΔInc.*R*^2^, median percent change in average Inc.*R*^2^ across clusters; *age and sex among proxy cases in the proxy analysis are the age and sex of the affected parent or average of affected parents.

We next aimed to prioritize the inclusion of SNPs in open chromatin regions, hypothesizing that these regions are more likely to play a role in pathogenesis. We applied a hybrid C+T approach with the *p-*value threshold applied only to SNPs in closed chromatin regions^37^. When the hybrid C+T method was applied to all annotation strategies, the S-3 annotation again yielded pathway-PRS with stronger predictive performance (median relative difference: 28.3% [IQR:-0.8%, 48.0%]; **Fig. 5b**). This pattern was consistent across analyses (**Supplementary Figs. 9-11**). While S-3 pathway-PRS performed best across C+T methods, the hybrid method performed worse than standard C+T overall (**Supplementary Fig. 12**).

As Hi-C data is not publicly available for all tissue and cell types, we tested a modification of the S-3 annotation strategy deriving annotation of SNPs to their regulatory targets using enhancer-gene pairs predicted by the Activity-by-Contact (ABC) model^19,38^. We performed this annotation based on publicly available predictions for microglia, astrocytes and bipolar neurons^19^. ABC-derived annotations did not improve pathway-PRS over S-1 annotations as did S-3, potentially due to the reduction in SNPs annotated to clusters from narrower enhancer-region boundaries in ABC (**Supplementary Figs. 13-17**).

### Improved detection of AD, sex-dependent, and age-at-onset associations

To test the replicability of our findings we calculated best-fit PRS trained in the UKB proxy data on 3,370 ADGC participants (**Table 1**; see Methods). As in the UKB, several pathway-PRS outperformed the global PRS (**Fig. 6a**). The performance gain with S-3 annotation in the ADGC was greater than that observed in the UKB (median relative difference:86.5% [IQR:0%, 204.8%]), likely due to greater precision of the AD outcome in the ADGC. The number of AD risk loci represented in pathway-PRS positively correlated with performance (**Fig. 6b; Supplementary Table 1**) but did not fully explain the differences between S-1 and S-3. In summary, our primary finding showing improved pathway-PRS performance under integrative annotation was replicated in the ADGC.

**Figure 6.**
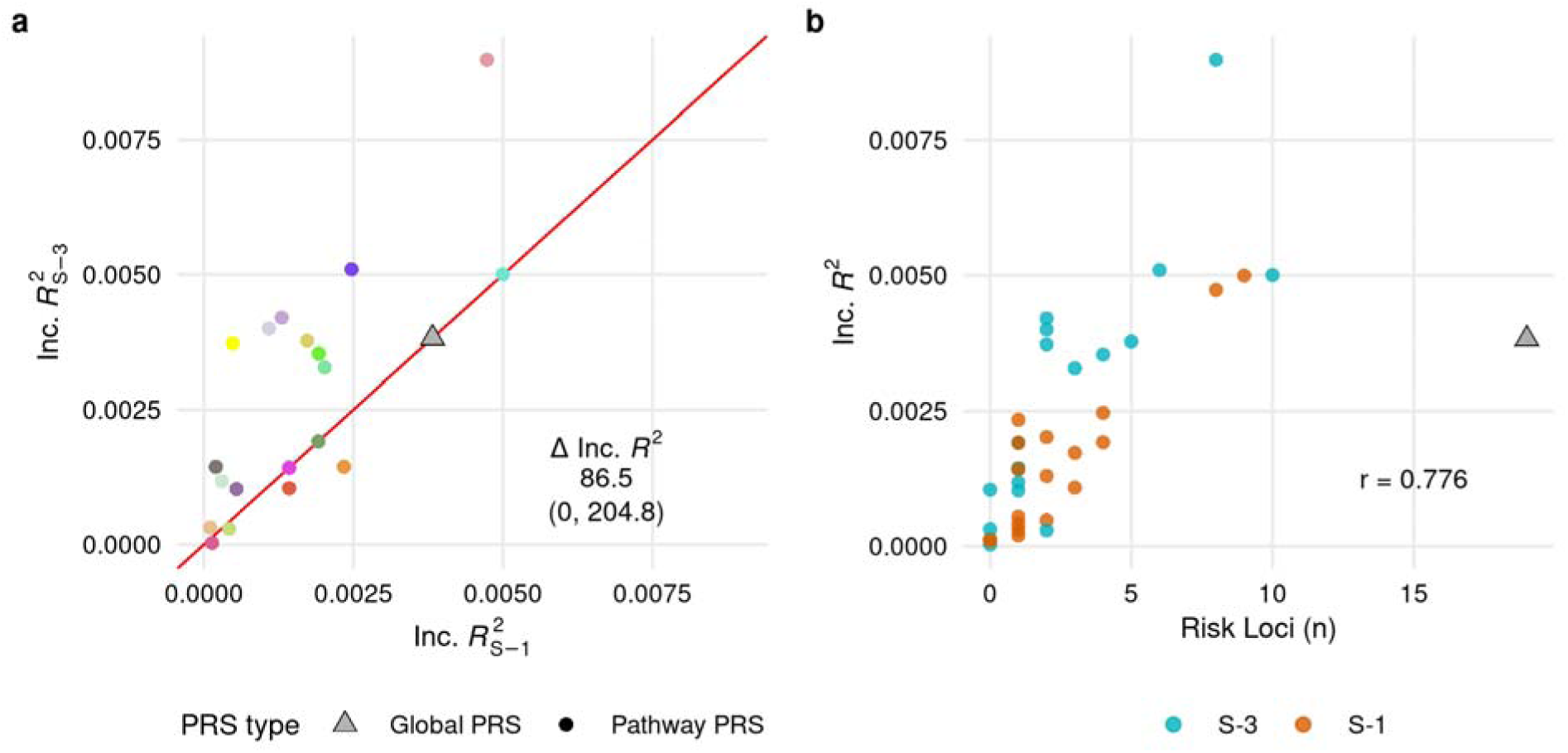
Pathway-PRS performance in the ADGC by annotation strategy and represented risk loci. Global and pathway-PRS trained on the UK Biobank proxy analysis dataset and calculated in the ADGC; **(a)** predictive performance across S-1 and S-3 annotation, and **(b)** number of genome-wide significant AD risk loci represented in the PRS model compared to predictive performance. Inc.*R_X_*^2^, incremental *R*^2^ for each PRS as full model compared to covariate-only model; ΔInc.*R*^2^, percent change in Inc.*R*^2^ as median (Q1, Q3); r, spearman correlation between number of risk loci and incremental *R*^2^; Genome-wide significant risk loci obtained from Kunkle et al 2019 (Supplementary Table 8).

Odds ratios (ORs) were estimated for PRS associations with AD in the ADGC. S-3 pathway-PRS models tended to include more SNPs and show stronger associations with AD risk than S-1 models (**Fig. 7**). Of pathway-PRS with consistent AD-associations across annotation strategies two were significantly associated at the corrected threshold (*P<*0.0025; C3, C5); five were significantly associated at the nominal threshold (*P<*0.05; C2, C7, C14, C15, C18); and eight were not associated. Among pathway-PRS with consistent significant associations with AD, the magnitude of associations under S-3 were stronger (4 pathway-PRS) or equal (3 pathway-PRS) to those under S-1 (**Supplementary Table 2**). Finally, five pathway-PRS were associated with AD dependent on annotation strategy with three significantly associated only under S-3 (*P<*0.0025: C4, C9; *P<*0.05: C8), one significantly associated only under S-1/S-2 (*P<*0.05: C17), and one significantly associated at the corrected threshold under S-3 but only nominally significant under S-1 (C1).

**Figure 7.**
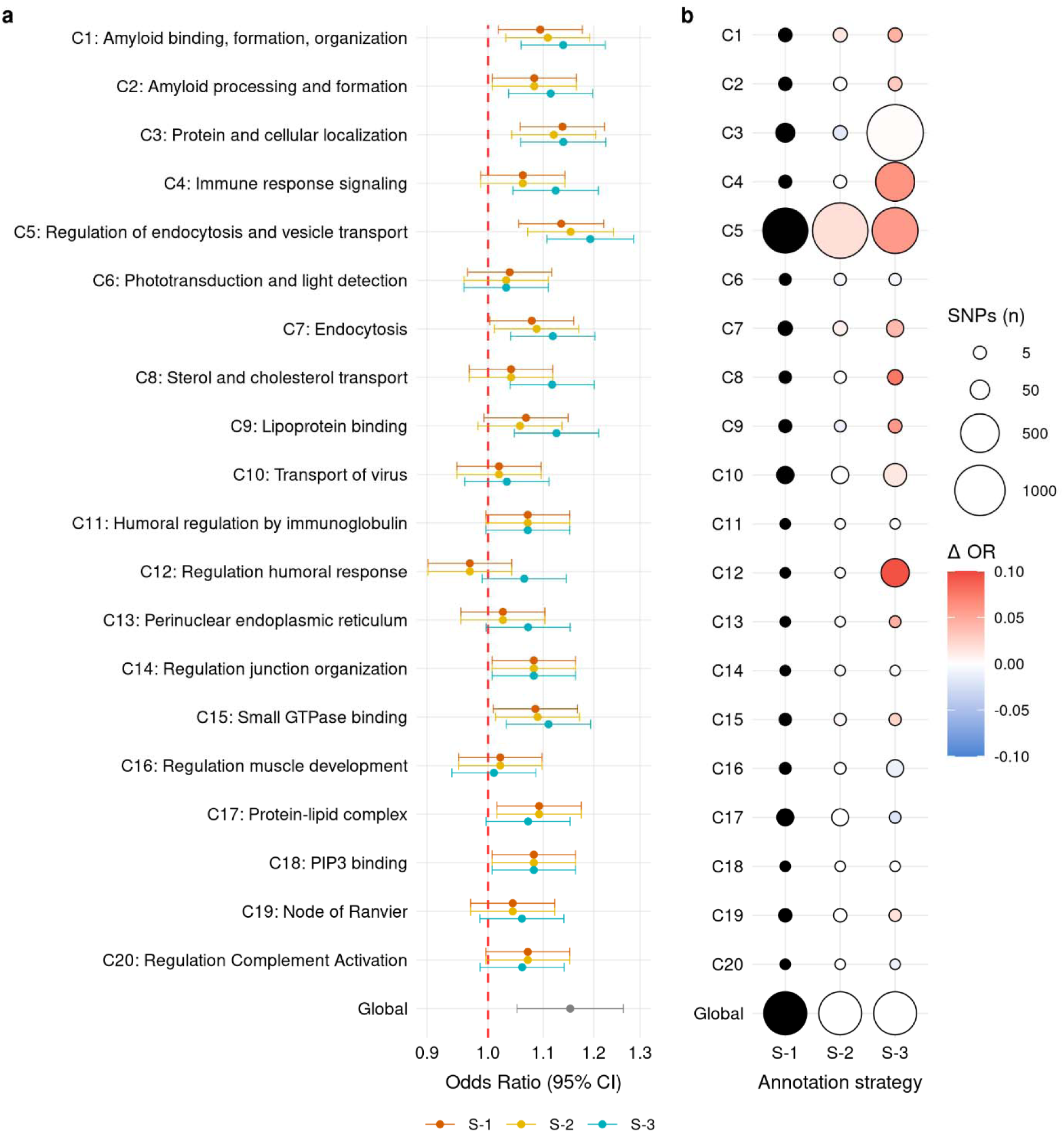
PRS size and association with AD in the ADGC. Global and pathway-PRS trained on the UK Biobank proxy analysis dataset and calculated in the ADGC; **(a)** association with AD status across annotation strategies and **(b)** size of PRS models as the number of included SNPs and relative strength of association with AD status across annotation strategies. ΔOR, difference in estimated odds ratio for the association with AD status from the S-1 pathway-PRS odds ratio.

The differences observed across annotation strategies affect inferences on the roles of various biological pathways. Under S-1, we would conclude that genetic risk in *lipoprotein binding* pathways (C9) may not have a role in AD pathogenesis independent of *APOE* but would come to the opposite conclusion under S-3 annotation. Additionally, under S-1, we would rank C3: *Protein and cellular localization* as the top associated pathway-PRS (OR=1.14, [95% CI:1.06, 1.22]), but under S-3 this would be C5: *Regulation of endocytosis and vesicle transport* (OR=1.19, [95% CI:1.11%, 1.29]; **Supplementary Table 2**). Findings were similar when pathway-PRS were trained on the true case data (**Supplementary Figs. 18-19; Supplementary Tables 3-4**).

We next sought to determine whether annotation improvements translate to improved detection of age-at-onset and sex-dependent associations. We regressed age-at-onset on pathway-PRS with covariate adjustment in the ADGC cases (n = 1,489 with age-at-onset). A single pathway-PRS, C3: *Protein and cellular localization*, was nominally associated with earlier onset when constructed under S-3 (Beta= -0.48, *P*=5.89×10^-03^; **Supplementary Table 5**). In a sex-stratified analysis, one pathway-PRS, C5: *Regulation of endocytosis and vesicle transport*, was significantly associated with AD in females at the corrected threshold under both S-1 (OR=1.22, *P*=6.89×10^-04^) and S-3 (OR=1.29, *P*=3.19×10^-05^), but not in males under S-1 (OR=1.01, *P*=0.85) or S-3 (OR=1.07, *P*=0.22). Additionally, the pathway-PRS, C7: *Endocytosis*, met the corrected threshold in females under S-3 (OR=1.22, *P*=8.15×10^-04^) but not under S-1 (OR = 1.10, *P*=0.10) or in males under S-1 (1.07, *P*=0.25) or S-3 (OR=1.03, *P*=0.57; **Supplementary Table 6**). Results were similar when pathway-PRS were trained on true case data (**Supplementary Tables 7-8**). Thus, integrative SNP-to-gene annotation not only enhances overall pathway-PRS performance but also improves power to detect biologically interpretable genetic associations with stratified outcomes and endophenotypes.

## Discussion

Pathway-PRS have great potential to elucidate genetically driven mechanisms of disease. However, existing applications are limited by the implicit assumption that regulatory elements are located close to the genes they regulate. This assumption is often violated^18–20^. As demonstrated in this work, such violations can reduce the predictive performance, power, and interpretability of pathway-PRS. We performed a direct comparison of SNP-to-gene annotation strategies integrating varying levels of gene annotation and tissue-specific Hi-C and eQTL data to evaluate how integrative annotation influences pathway-PRS performance and the interpretation of their associations with AD. Our results support the use of functionally informed SNP-to-gene annotation in pathway-PRS construction and are relevant to complex traits beyond AD.

We observed the strongest predictive performance in pathway-PRS constructed using S-3, the most integrative SNP-to-gene annotation strategy. This result was consistent across sensitivity analyses, application of a hybrid C+T method, and replication in an independent dataset with rigorous ascertainment of the AD phenotype. Although S-3 annotated fewer SNPs to clusters due to fewer SNPs being captured by Hi-C than by the 35kb upstream and 10kb downstream boundaries, it included more SNPs in the majority of trained pathway-PRS models, suggesting greater annotation precision. While S-3 models typically included more SNPs than S-1/S-2 models, several S-3 models of equal or smaller size to S-1 models achieved greater predictive performance, suggesting that SNPs annotated by S-3 are more relevant to AD. This improvement cannot be explained by differences in the number of known genome-wide significant AD loci represented in each pathway-PRS, as represented risk loci remained stable across annotation strategies for several clusters despite improved performance (**Supplementary Tables 3-4**). Notably, several S-3 pathway-PRS outperformed the global PRS. This finding challenges the expectation that pathway-PRS are inherently less predictive than global PRS and is consistent with performance gains observed in global PRS under other integrative methods, such as LDpred-funct^39^ and SBayesRC^40^.

The S-3 annotation strategy adapts and extends the *H-MAGMA* integrative framework to the pathway-PRS context, including eQTLs in addition to Hi-C and gene annotation. Several prior studies have incorporated functional data into pathway-PRS construction^4,13,41,42^, though few have compared performance across annotation strategies. Pistis et al. observed stronger associations between pathway-PRS and psychosis using pathway-PRS based only on eQTLs as compared to positional annotation^43^. Cell-type- and cell-state-specific PRS may also be constructed by annotating SNPs to gene sets, thus effects of functional data integration may be expected to generalize across PRS frameworks. Consistent with this, Yang et al. found no difference in trait associations with a cell-type-specific PRS based on a positional strategy with varying window sizes, whereas Ord et al.^37^ reported cell-state-specific PRS based on single-nucleus chromatin accessibility data to perform better than those based on positional annotation. The S-3 strategy presented here integrates bulk-tissue derived eQTLs and Hi-C data, rather than single-cell or single-nucleus assays. We observed little benefit from the integration of cell-type-specific ABC enhancer-gene pairs, though this may be due to a reduction in the number of SNPs annotated with the ABC-derived enhancer-gene pairs rather than a limitation of cell-type-specific annotation. While cell-type-specific chromatin interaction data can be underpowered, it enables the investigation of refined biological hypotheses, and further work in this area is warranted.

There are several limitations to our analysis. First, publicly available functional data are largely generated on post-mortem samples^20^ obtained primarily from individuals with genetic similarity to European reference populations^23^. Though there is some evidence that these data are moderately transferable across tissues of living and deceased donors^44–46^ and across populations^23,47,48^ the extent of transferability is not fully understood. In future work, we will evaluate S-3 across more complex methods, such as PRS-CS^49^, and across populations using multi-ancestry PRS methods^50^.

Second, additional functional data modalities, such as chromatin or transcript usage QTLs^13^ could further improve pathway-PRS performance. In this work, we limited functional data to types consistently available across tissues to maximize the flexibility of the S-3 strategy^23,51^. Finally, we selected Gene Ontology terms based on a pathway enrichment analysis using positional SNP-to-gene annotation^32^. This aligns with prior pathway enrichment analyses^29,52^. As a result, the selected pathways are enriched for GWAS signal under S-1 and comparisons of annotation strategies are biased in favor of S-1. Therefore, the performance gains we observed are likely conservative estimates of the benefit of S-3 on pathways with known relevance to AD. In fact, despite the reduced GWAS-significance of SNPs annotated under S-3 as compared to S-1, S-3 improved the performance of the majority of pathway-PRS. The few pathway-PRS with decreased performance under S-3 may indicate that S-3 failed to capture their AD-relevant pathway-SNPs. Alternatively, the S-3 strategy may be accurately resolving an annotation which misattributed AD-relevance to these pathways, consistent with their exclusion from prior AD pathway literature^29,53^.

The most integrative SNP-to-gene annotation strategy, S-3, captures SNPs with greater relevance to tissue-specific gene function and consistently improves pathway-PRS performance. These gains highlight the importance of long-range gene regulatory relationships in AD pathogenesis. Notably, several S-3 pathway-PRS outperformed the global PRS, underscoring the value of prioritizing functionally relevant SNPs over simply increasing SNP quantity and alleviating concerns of reduced statistical power in pathway-as compared to global-PRS analyses. While these findings are encouraging, we acknowledge the limitations of the S-3 strategy and recommend the application of multiple SNP-to-gene annotation strategies to support robust inference, along with clear and detailed reporting of annotation strategies to ensure reproducibility. S-3 builds on the *H-MAGMA* framework, which is straightforward to implement and leverages genome-wide SNP-to-gene annotation, making it compatible with many PRS construction methods and enabling broad adoption of integrative annotation across pathway-PRS and related PRS frameworks that rely on annotation of SNPs to gene sets.

## Methods

### Data sources

European genetic ancestry participants from the UKB^26^ were used for pathway-PRS training and testing (**Supplementary Figs. 1-3**). We used independent, ADGC European genetic ancestry samples from the Alzheimer’s Disease Centers (ADCs) waves 8-12 for replication (n = 3,370; see supplementary methods).

Summary statistics for pathway enrichment analysis and pathway-PRS construction were obtained from the 2019 IGAP GWAS^29^, which did not include participants from the UKB or ADC waves 8-12. The 1000 Genomes Project Phase 3^54^ European superpopulation was used as an LD reference panel.

### Pathway Enrichment Analysis and Pathway Clustering

We selected pathways with relevance to AD though *MAGMA*^32^ pathway enrichment analysis of AD GWAS signal in GO pathways as in Schork et al. 2023^17^. IGAP summary statistics^29^ were filtered to exclude duplicate and ambiguous SNPs and the *APOE* region (Chr19:43,873,587-45,621,890, hg38) to minimize prioritization of pathways based on a single gene^52^. We annotated SNPs to protein-coding genes based on position using a window 35kb upstream (5’) and 10kb downstream (3’) of gene boundaries. Pathway annotations were derived from the Bader Lab website (http://baderlab.org/GeneSets, downloaded May 1, 2024) and included GO annotations with at least one evidence code other than IEA (Inferred from Electronic Annotation) containing 10-1,000 annotated genes^17,55,56^. The competitive analysis was run using the SNP-wise mean model. We retained pathways meeting FDR<0.25 following Benjamini-Hochberg correction, a permissive threshold selected to include a broad range of pathways. Pathways were collapsed into clusters based on overlapping gene content with *EnrichmentMap*-v3.3.6^57^ to minimize redundancy (overlap threshold = 0.4) and *AutoAnnotate*-v1.4.1^58^ (MCL algorithm) in *Cytoscape*-v3.10.1^59^.

### Ascertainment of AD Status

In the proxy analysis we defined AD cases as true or proxy cases. True case status was defined by the presence of at least one of a validated set of ICD-9/10 codes for AD in inpatient records or death certificates^60^. Participants who did not meet true case criteria, did not have ICD-9/10 codes for other dementias^60^, and were not adopted were eligible to be proxy cases or controls. Proxy case status was defined by positive report of maternal and/or paternal AD/dementia at baseline or follow up assessment^61^. Controls were further restricted to those of 50+ years of age at data freeze.

In the true case analysis, we restricted all participants to those older than 50 years at baseline assessment. Cases included only those with ICD-9/10 codes for AD in inpatient records or death certificates and with age of onset over 65 years (criteria for late-onset AD; **Supplementary Fig. 2**). Controls were those with no other dementia diagnosis^60^. The Wu/Marioni sensitivity analysis implemented the methods of Marioni et al. 2018 and Wu et al. 2024 shown to partially correct for survival bias^30,31^. This analysis did not assess true case status and excluded individuals who were adopted or missing adoption status. Proxy cases were those who reported at least one parent with AD/dementia. Controls were those with both parents unaffected. If parental age was under 65 years at parental death or time of participant assessment then parental AD/dementia status was considered missing (**Supplementary Fig. 3**).

AD status in the ADGC was defined as meeting clinical criteria for possible or probable AD^62,63^. Controls were cognitively normal elders aged 60 years or older with MMSE scores >28 or reported cognitively intact at time of death (see Supplementary Methods).

### UKB covariate definitions

In the proxy and true case analyses, we adjusted for age, sex, number of *APOE ε*4 alleles, and the top five principal components (PCs) in our global and pathway-PRS models. In the proxy analyses, proxy case age and sex were defined based on the average of affected parent values at time of parental death or participant report. For controls and true cases, sex was defined as participant self-reported sex and age was defined as age of AD onset in years, if available, or age at death or data freeze.

The Wu/Marioni sensitivity analysis included the same covariates used by Wu et al. 2024^30^. This included participant year of birth, sex as participant self-reported sex, number of *APOE ε*4 alleles, top five PCs, genotyping batch and both father’s and mother’s age at first assessment if available or at death.

### SNP-to-gene annotation

IGAP summary statistics were filtered to exclude ambiguous or duplicated SNPs and all SNPs in the *APOE* region (Chr19:43,873,587-45,621,890, hg38) prior to annotation. The S-1 SNP-to-gene annotation strategy was implemented in *MAGMA*^32^ and included SNPs within 35 kilobase pairs (kb) upstream and 10 kb downstream of NCBI 37.3 boundaries for protein-coding genes (data from the Complex Trait Genetics website; https://cncr.nl/research/magma/). For S-2 annotation, the IGAP summary statistics were run through the *FUMA*^34^ SNP2GENE pipeline with annotation of SNPs in LD (*r^2^*≥0.6) with a lead SNP (*P*<10^-5^, LD-independent at *r^2^*=0.6) using eQTL and 3D chromatin interaction mapping with PsychENCODE eQTL and promoter anchored loops^34^. The S-3 annotation strategy was implemented as in Sey et al. 2023^35^ which captures SNPs in GENCODE v26 exons and promoter regions (2kb upstream of the transcription start site) and SNPs in regions observed to contact the promoter region in adult brain Hi-C data. Exon, promoter, and Hi-C data were obtained from the H-MAGMA protocol, with Hi-C data derived from PsychENCODE adult brain tissue (n_brains_=3; https://zenodo.org/records/6382668)^35,36^. In addition to the Sey protocol for S-3, we annotated eQTL SNPs to their target genes using significant eQTLs in adult brain tissue (FDR<0.05) obtained from PsychENCODE (n_brains_=1,387; http://resource.psychencode.org/)^20^. Annotated SNP sets were filtered to exclude those with missing genotype frequency>0.01, with a minor allele frequency (MAF)<0.01, or an imputation INFO score of <0.8 in the UKB.

The S-3 annotation strategy was additionally adapted to use ABC predicted enhancer-gene pairs in place of PsychENCODE Hi-C data. ABC predictions for microglia were obtained from the Fresh Microglia Study. Predictions for astrocytes and bipolar neurons were obtained from https://www.engreitzlab.org/resources/^19^. Predicted pairs were filtered to those with ABC score≥0.02. SNPs within the predicted enhancer bounds were annotated to the paired gene. We also applied ABC-Max by restricting predicted enhancer-gene pairs to those with the highest ABC Score for a given enhancer^19^.

### PRS Construction and Evaluation

In each of 10 repeated holdouts, the UKB data was split 80:20 using random splitting stratified by AD case/proxy/control status and sex. In the 80% training sets, we applied C+T on annotated SNPs using default parameters (1 Mb window size, *r^2^*=0.1, *P*=1) and seven *p*-value thresholds (5×10^-8^, 5×10^-6^, 5×10^-4^, 0.05, 0.1, 0.5, and 1) in *PRSet*^7^, allowing for the inclusion of LD-proxies (*r^2^*≥0.8) in determining index SNPs.

Best-fit pathway-PRS across *p-*value thresholds had the highest incremental *R^2^* values for the full compared to covariate-only models. Global and pathway-PRS were constructed as a standardized weighted sum of risk alleles with SNP weights derived from the IGAP GWAS^29^.

We additionally applied a hybrid C+T approach with the *p-*value threshold applied only to SNPs in closed chromatin regions^37^. Chromatin state data were obtained for adult brain dorsolateral pre-frontal cortex from the Roadmap Epigenomics Project core 15-state model^64,65^. States 1-7 were considered open chromatin and all others closed chromatin.

In the testing sets, AD status was regressed on covariates and best-fit pathway-PRS. Pathway-PRS performance across annotation strategies was measured as the average of testing set incremental *R^2^*values across repeated holdouts.

### Principal component analysis in the ADGC

PCs for population stratification adjustment were estimated in *PLINK*-v2.0^66^. We pooled data across ADCs 8-12, filtered genotyped SNPs with missing call rates>0.1, MAF<0.01, and departure from Hardy-Weinberg Equilibrium (HWE) at *P*<10^-6^. We pruned the filtered genotype data with a window size of 1000 SNPs and step size of 250, filtering SNPs in LD (*r^2^*>0.1). We verified that no related individuals up to the 3^rd^ degree remained in the data using KING-robust in *PLINK*-v2.0^66^ before computing loadings for 10 PCs. The top three PCs were selected as covariates from examination of a scree plot.

### Replication of pathway-PRS performance in the ADGC

Annotated SNPs were filtered to exclude those with MAF<0.01, INFO<0.5, HWE *P*<10^-6^, and missing call rate>0.1 in the ADGC data. We trained PRS in the UKB on the full proxy and true case datasets using C+T in *PRSet*^7^, as above. We used *rtracklayer*-v1.64.0^67^ to lift over pathway-PRS models from GRCh37 to GRCh38 before calculating best-fit PRS in the ADGC. Dosages for missing SNPs were imputed as the SNP MAF in the 1000 Genomes Project EUR superpopulation LD reference. We regressed AD status on PRS adjusted for age, sex, number of *APOE ε*4 alleles, and PCs. We assessed pathway-PRS predictive performance with incremental *R^2^* for the full as compared to covariate-only models. ORs for the association of pathway-PRS with AD were estimated for a 1 SD change in PRS. Age-at-onset analyses included all covariates except age^68,69^. Sex-stratified analyses included adjustment for all covariates except sex. In the sex-stratified analyses, female cases and controls were randomly sampled such that male and female analyses were equally powered (cases=729, controls=683). In all tests, *P*<0.05 was considered nominally significant, *P*<0.0025 was considered significant given Bonferroni correction for 20 independent tests. Analyses were conducted with *R*-v4.3^70^.

### AI use

The authors acknowledge the use of ChatGPT (GPT-4 and GPT-5; OpenAI), accessed throughout the research process from October 2024 to May 2026 to assist with debugging and improving code. All AI generated code was carefully reviewed and revised by the authors, and the final content reflects their contributions.

## Data availability

Data needed to generate main figures and SNPs included in the global and pathway-PRS in the ADGC replication are available in the supplementary materials. Weights for included SNPs from the IGAP GWAS can be obtained as described below. Other summary-level data produced in the present study are available upon reasonable request to the authors.

The ADGC datasets used in this study are available through the NIAGADS Data Sharing Service at dss.niagads.org under accession numbers NG00136, NG00137, NG00138, NG00139, and NG00140.

IGAP GWAS summary statistics are available at https://dss.niagads.org/datasets/ng00075/.

The UK Biobank data used in this study were available under UK Biobank approval (application 50806). UK Biobank data are available through the UK Biobank (http://www.ukbiobank.ac.uk/) upon application and with permission of UKBB’s Research Ethics Committee.

LD-reference and gene boundaries for MAGMA pathway enrichment analyses are available at https://cncr.nl/research/magma/.

The code and materials used for SNP-to-gene annotation with the H-MAGMA pipeline are available at https://zenodo.org/records/6382668.

The PsychENCODE functional genomics data is available at http://resource.psychencode.org/.

Gene Ontology pathway annotations are available at http://baderlab.org/GeneSets.

Roadmap Epigenomics chromatin state data are available at https://egg2.wustl.edu/roadmap/data/byFileType/chromhmmSegmentations/ChmmModels/coreMarks/jointModel/final/.

ABC predicted enhancer-gene pairs are available at https://www.engreitzlab.org/resources/.

The Fresh Microglia Regulome data, analyses and tools are shared early in the research cycle without a publication embargo on secondary use. Data is available for general research use according to the following requirements for data access and data attribution. For access to predicted enhancer-gene pairs in microglia see: http://doi.org/10.7303/syn26207321.

## Code availability

Code for the primary analyses can be downloaded from https://github.com/KSBazemore/pathPRS_by_annotation.git.

## Acknowledgements

We thank the participants of the UK Biobank for their invaluable contributions to this research.

We thank the International Genomics of Alzheimer’s Project (IGAP) for providing summary results data from the IGAP Rare Variant GWAS (Kunkle et al., 2019) for these analyses. The IGAP investigators provided data but did not participate in design, analysis, or writing of this report. IGAP was made possible by the generous participation of the control subjects, the patients, and their families.

Samples from the National Centralized Repository for Alzheimer’s Disease and Related Dementias (NCRAD), U24 AG021886, were used in this study. The NACC database is funded by NIA/NIH Grant U24 AG072122. NACC data are contributed by the NIA-funded ADRCs: P30 AG062429 (PI James Brewer, MD, PhD), P30 AG066468 (PI Oscar Lopez, MD), P30 AG062421 (PI Bradley Hyman, MD, PhD), P30 AG066509 (PI Thomas Grabowski, MD), P30 AG066514 (PI Mary Sano, PhD), P30 AG066530 (PI Helena Chui, MD), P30 AG066507 (PI Marilyn Albert, PhD), P30 AG066444 (PI John Morris, MD), P30 AG066518 (PI Jeffrey Kaye, MD), P30 AG066512 (PI Thomas Wisniewski, MD), P30 AG066462 (PI Scott Small, MD), P30 AG072979 (PI David Wolk, MD), P30 AG072972 (PI Charles DeCarli, MD), P30 AG072976 (PI Andrew Saykin, PsyD), P30 AG072975 (PI David Bennett, MD), P30 AG072978 (PI Neil Kowall, MD), P30 AG072977 (PI Robert Vassar, PhD), P30 AG066519 (PI Frank LaFerla, PhD), P30 AG062677 (PI Ronald Petersen, MD, PhD), P30 AG079280 (PI Eric Reiman, MD), P30 AG062422 (PI Gil Rabinovici, MD), P30 AG066511 (PI Allan Levey, MD, PhD), P30 AG072946 (PI Linda Van Eldik, PhD), P30 AG062715 (PI Sanjay Asthana, MD, FRCP), P30 AG072973 (PI Russell Swerdlow, MD), P30 AG066506 (PI Todd Golde, MD, PhD), P30 AG066508 (PI Stephen Strittmatter, MD, PhD), P30 AG066515 (PI Victor Henderson, MD, MS), P30 AG072947 (PI Suzanne Craft, PhD), P30 AG072931 (PI Henry Paulson, MD, PhD), P30 AG066546 (PI Sudha Seshadri, MD), P20 AG068024 (PI Erik Roberson, MD, PhD), P20 AG068053 (PI Justin Miller, PhD), P20 AG068077 (PI Gary Rosenberg, MD), P20 AG068082 (PI Angela Jefferson, PhD), P30 AG072958 (PI Heather Whitson, MD), P30 AG072959 (PI James Leverenz, MD).

The results published here are in part based on data from the Fresh Microglia Regulome Study obtained from the AD Knowledge Portal (https://adknowledgeportal.org/). We thank the patients and families who donated material for these studies. We thank the computational resources and staff expertise provided by the Scientific Computing group at the Icahn School of Medicine at Mount Sinai.

## Funding Statement

K.B. discloses support for the research of this work from the National Institute on Aging (NIA; F31AG09198). A.C.N., T.I., A.B.K., G.D.S., and L-S.W. disclose support for the research of this work from the NIA-supported ADGC (U01AG032987 and RC2AG036528), the NIA Genomics of Alzheimer’s Disease Data Storage Site (NIAGADS; U24AG041689), and the NIA-supported Genome Center for Alzheimer’s Disease (GCAD; U54AG052427). J.J. discloses support for the research of this work from the National Human Genome Research Institute (R00HG012223), the National Institute of General Medical Sciences (R35GM157133), and the Penn Center for Eye-Brain Health through the CCEB Innovation Award. S.F.A.G. and A.C. declare no relevant funding.

## Author Contributions

K.B., A.C.N, and J.J. conceived and designed the study. S.F.A.G., L-S.W., and A.C., gave instrumental feedback on the study design. K.B. performed analyses, T.I. assisted with analyses. K.B. and A.C.N. wrote the manuscript with the participation of all authors. A.B.K., G.D.S., and L-S.W. contributed data. All authors reviewed and approved the final manuscript.

## Ethics Statement

Ethics approval for the UK Biobank study was obtained from the North West Centre for Research Ethics Committee (11/NW/0382). The ADGC protocol is reviewed and approved by the University of Pennsylvania IRB #8 (Federalwide Assurance #00004028). Informed consent was obtained from all human research participants.

## Competing Interest Statement

All authors have completed the ICMJE uniform disclosure form at www.icmje.org/coi_disclosure.pdf and declare: L-S.W. received payment/honoraria for lectures at Academia Sinica, Taipei, Taiwan, China Medical University, Taichung, Taiwan, Taichung Veterans General Hospital, Taiwan, and University of Michigan; A.C.N received honoraria for speaking at a seminar from the American Statistical Association, Columbus Ohio Chapter; S.F.A.G. receives funding from the Daniel B. Burke Endowed Chair for Diabetes Research at CHOP; K.B., A.B.K., G.D.S., L-S.W., J.J., and A.C.N. have received grant funding from the National Institutes of Health, including from the National Institute on Aging and others; no other relationships or activities that could appear to have influenced the submitted work.

## Notes

### Author Declarations

Ethics approval for the UK Biobank study was obtained from the North West Centre for Research Ethics Committee (11/NW/0382). The ADGC protocol is reviewed and approved by the University of Pennsylvania IRB #8 (Federalwide Assurance #00004028).

### Summary of Updates

Minor changes to language and figure captions

