## Supplementary Note for "Functionally informed annotation influences pathway-specific polygenic risk and disease inference in Alzheimer’s disease"

**Contents**

| Supplementary Methods | 2 |
| --- | --- |
| Supplementary Figures | 3 |
| References | 19 |

### Supplementary Methods

##### The UK Biobank Cohort

The UK Biobank cohort, including enrollment, phenotyping, genotyping, quality control, and imputation have been described in detail elsewhere^1^. Inclusion criteria for all analyses are detailed in Figures S1-3. Briefly, we excluded withdrawals, individuals with non-European genetic ancestry (UK Biobank data field 22006-0.0), those with genetic sex (field 22001-0.0) different to self-reported sex (field 31-0.0) or sex chromosome aneuploidy (field 22019-0.0), outliers for heterozygosity or missingness (field 22027-0.0), and related individuals up to the 3^rd^ degree (field 22020-0.0). Alzheimer’s Disease status was defined as true AD or proxy AD. True AD was determined by the presence of at least one of a previously validated set of ICD-9/10 codes for AD as primary/secondary diagnoses in inpatient records or as primary/secondary causes of death in death certificates (field 42021-0.0)^2^. Age at AD onset was calculated based on the data of Alzheimer’s disease report (field 42020-0.0) and participant month and year of birth (fields 52-0.0 and 34-0.0). Proxy AD was determined by participant report of parental AD/dementia in the family history report completed at the first assessment (fields 20107-0.[0-9] [father] and fields 20110-0.[0-9] [mother]). The top 5 principal components were obtained from the UK Biobank covariate data fields 22009-0.[1-5]. Number of *APOE* 𝜀4 alleles was inferred from genotypes at *APOE* haplotype tagging SNPs (rs7412 and rs429358). UK Biobank phenotype data for this analysis were downloaded on August 25, 2023; freeze of England hospital inpatient records prior to data download occurred on September 30, 2021. We included 328,536 individuals in the proxy analysis, 259,418 in the true case analysis, and 202,408 in the Wu/Marioni analysis.

##### Alzheimer’s Disease Genetics Consortium

The ADGC dataset includes participants from 4 waves of cases and cognitively normal controls from the National Institute on Aging (NIA) Alzheimer’s Disease Centers (ADCs). Data from ADC waves 8-12 were selected as they are independent of the waves included in the 2019 IGAP GWAS^3^. This data has been described in detail in prior ADGC studies^4^. In summary, the clinical and neuropathology cores at the NIA-funded ADCs enrolled and evaluated participants in the NIA ADC cohorts. Data was collected and cleaned by the National Alzheimer’s Coordinating Center (NACC). Alzheimer’s disease cases were classified as demented using NINCDS-ADRDA/DSMIV-V^5^ or NIA-AA criteria^6^ or by a Clinical Dementia Rating ≥ 137. We additionally excluded participants if they were analytical omits (poor genotyping quality, non-AD dementia cases, related individuals up to the 3^rd^ degree); did not meet AD case or control criteria; were duplicated samples across ADCs; or were less than 60 years of age. We defined cases as those meeting clinical criteria for possible or probable AD as described above. Controls were cognitively normal elders at least 60 years of age at last exam or death. Age-at-onset for AD cases was defined as previously described^7,8^. We included 3,370 individuals in total, 1,519 cases and 1,851 controls.

### Supplementary Figures

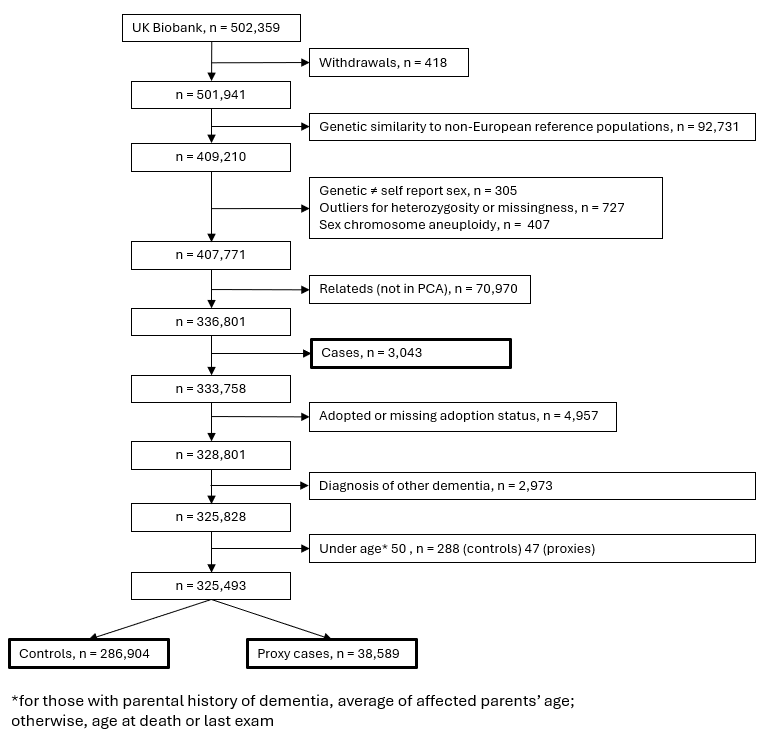

**Figure S1 |** **Proxy analysis inclusion flowchart**. UK Biobank participant inclusion flowchart, proxy analysis

*age in proxy cases is the average of affected parents’ age at death or at time of participant report

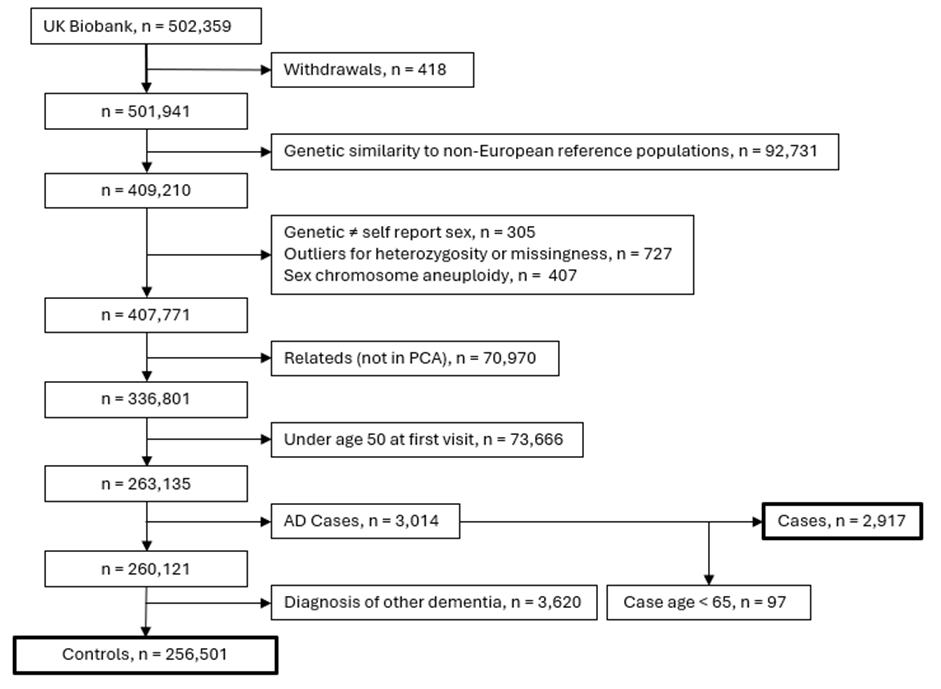

**Figure S2 |** **True case analysis inclusion flowchart**. UK Biobank participant inclusion flowchart, true case analysis

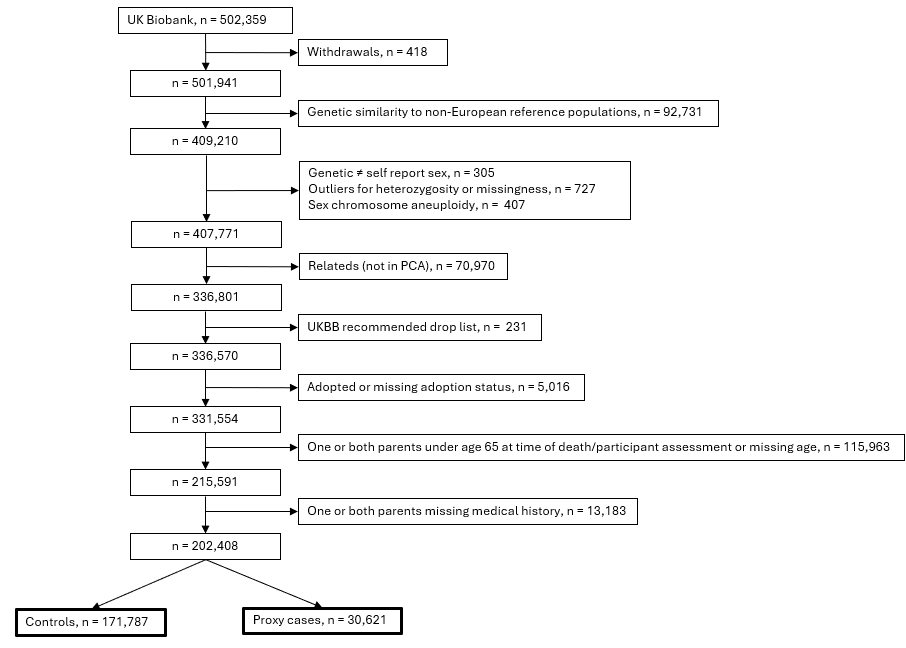

**Figure S3 |** **Wu/Marioni analysis inclusion flowchart**. UK Biobank participant inclusion flowchart, Wu/Marioni analysis

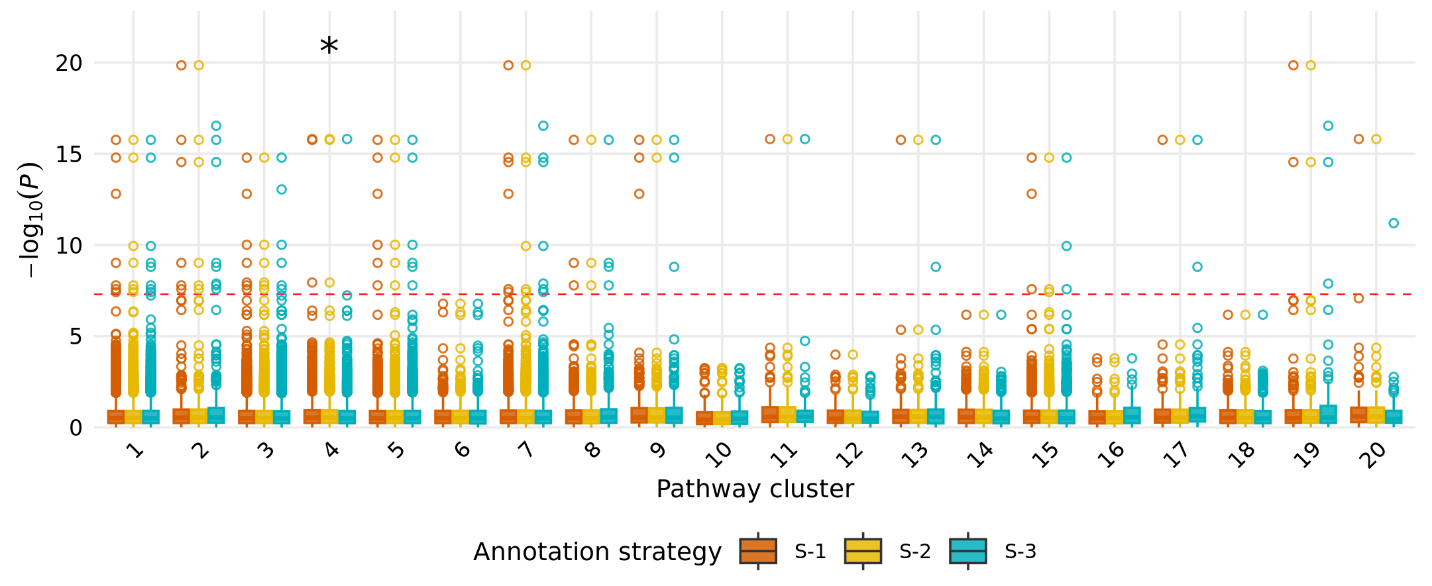

**Figure S4 |** ***P-*value distributions of LD-independent annotated SNPs across clusters by annotation strategy.** Kruskal-Wallis test ****p* ≤ 0.001, ***p* ≤ 0.01, **p* ≤ 0.05.

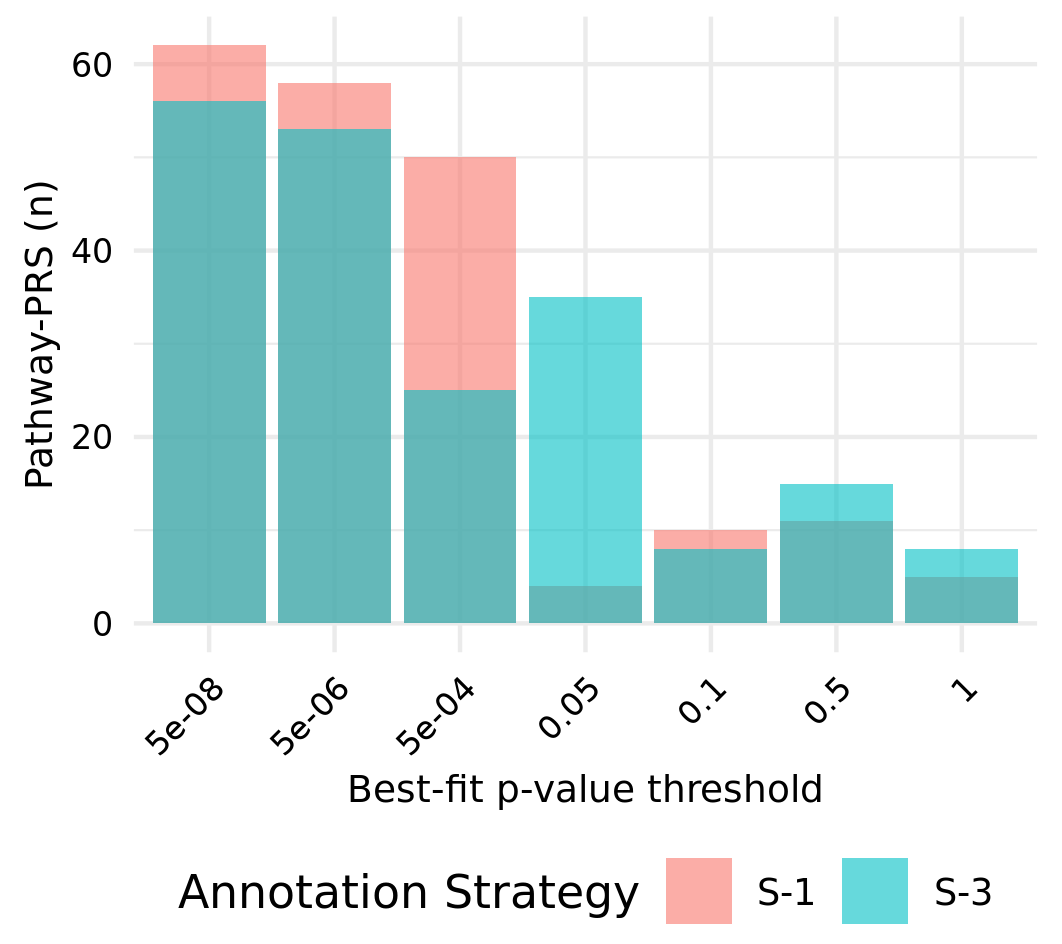

**Figure S5 |** **Pathway-PRS best-fit p-values**. Distributions of best-fit *p-*value thresholds of pathway-PRS by annotation strategy for all proxy analysis repeated holdouts.

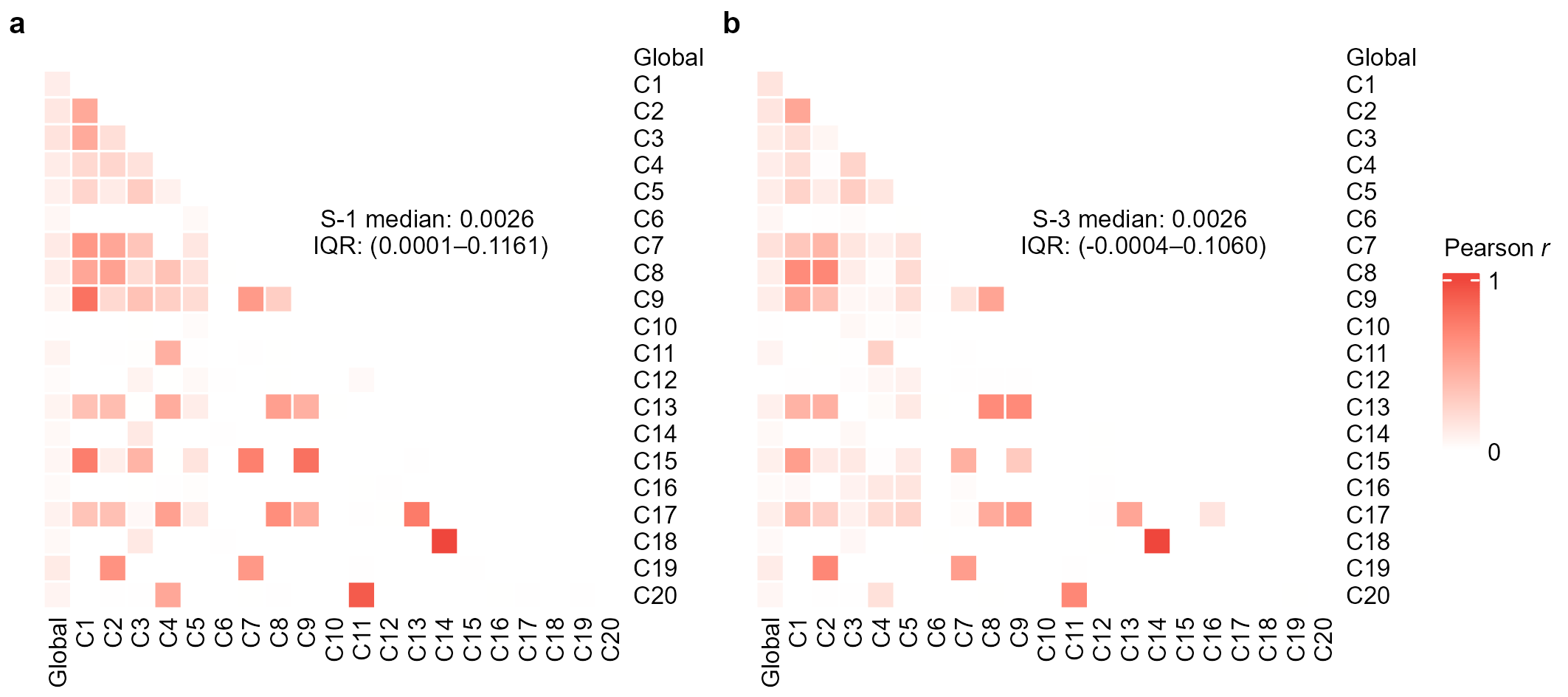

**Figure S6 |** **Pathway-PRS correlations.** Pearson correlations of calculated pathway-PRS under **(a)** S-1 annotation, and **(b)** S-3 annotation, each pairwise correlation is the median value across calculated correlations in each proxy analysis repeated holdout testing set.

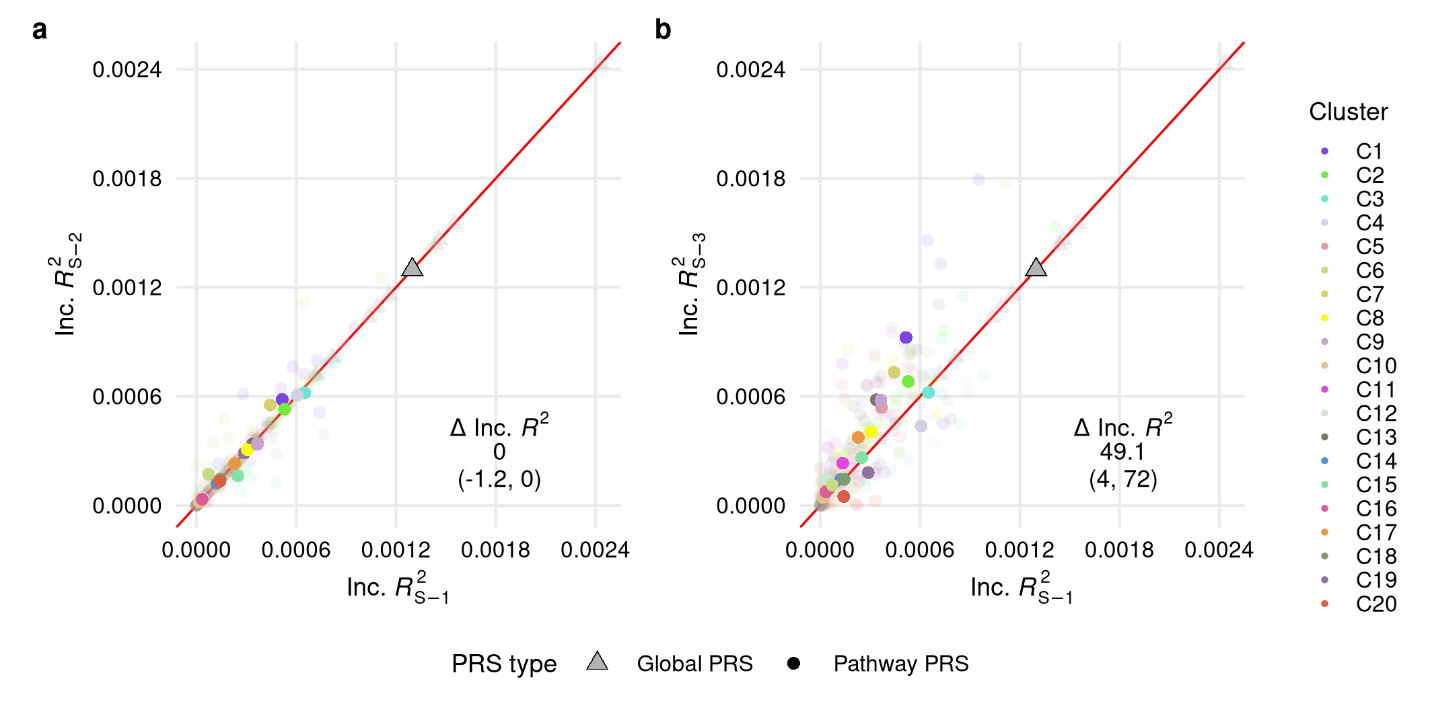

**Figure S7 | Wu/Marioni PRS performance.** Global and pathway-PRS predictive performance in the Wu/Marioni analysis UK Biobank testing sets under (a) S-1 vs S-2 annotation, (b) S-1 vs S-3 annotation.
Inc.$R_{X}^{2}$, incremental $R^{2}$ for each PRS as full model compared to covariate-only model; ∆Inc.$R^{2}$, percent change in Inc.$R^{2}$ as median (Q1, Q3); transparent points, Inc.$R^{2}$ estimates from individual holdouts; solid points, average Inc.$R^{2}$ over ten repeated holdouts.

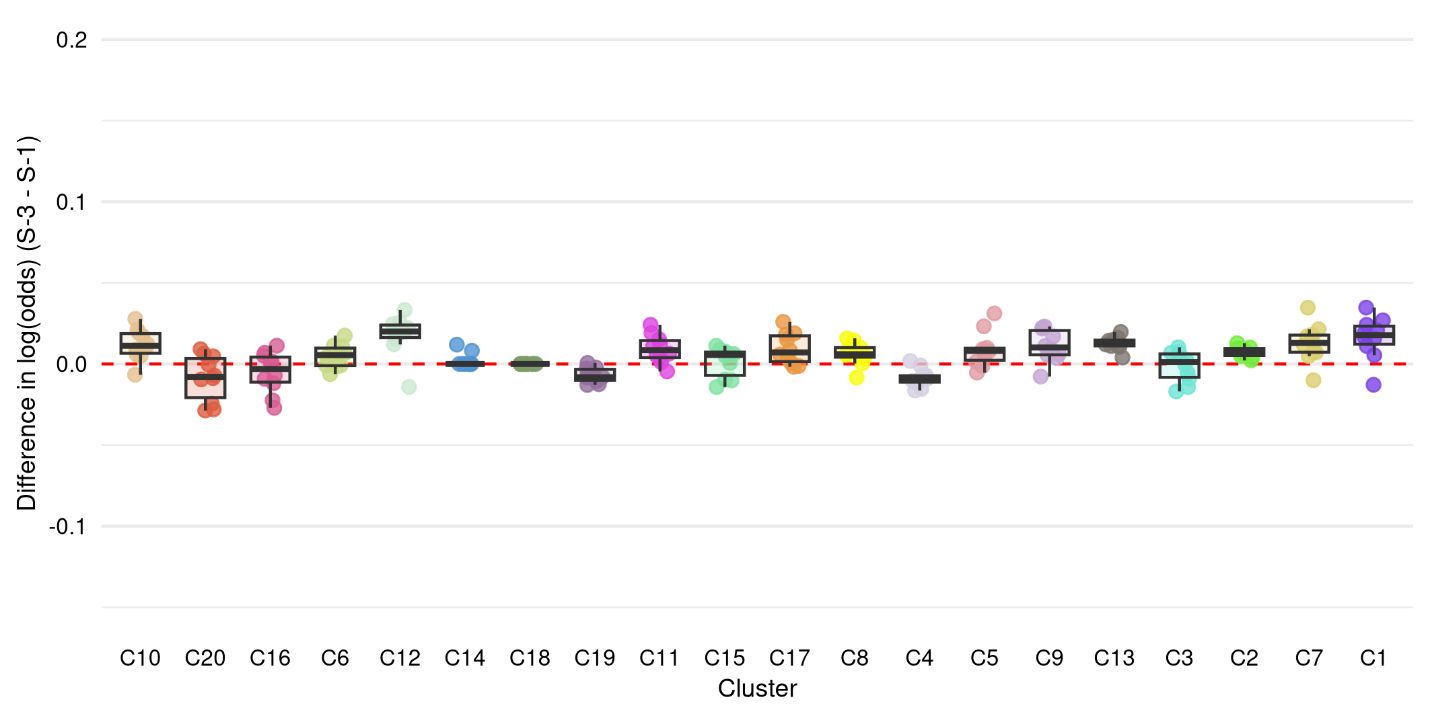

**Figure S8 |** **Change in magnitude of pathway-PRS association with AD in the Wu/Marioni analysis**. Change in association magnitude between S-1 and S-3 annotation strategies across repeated holdout testing sets adjusted for age, sex, number of *APOE* 𝜀4 alleles, and top PCs.

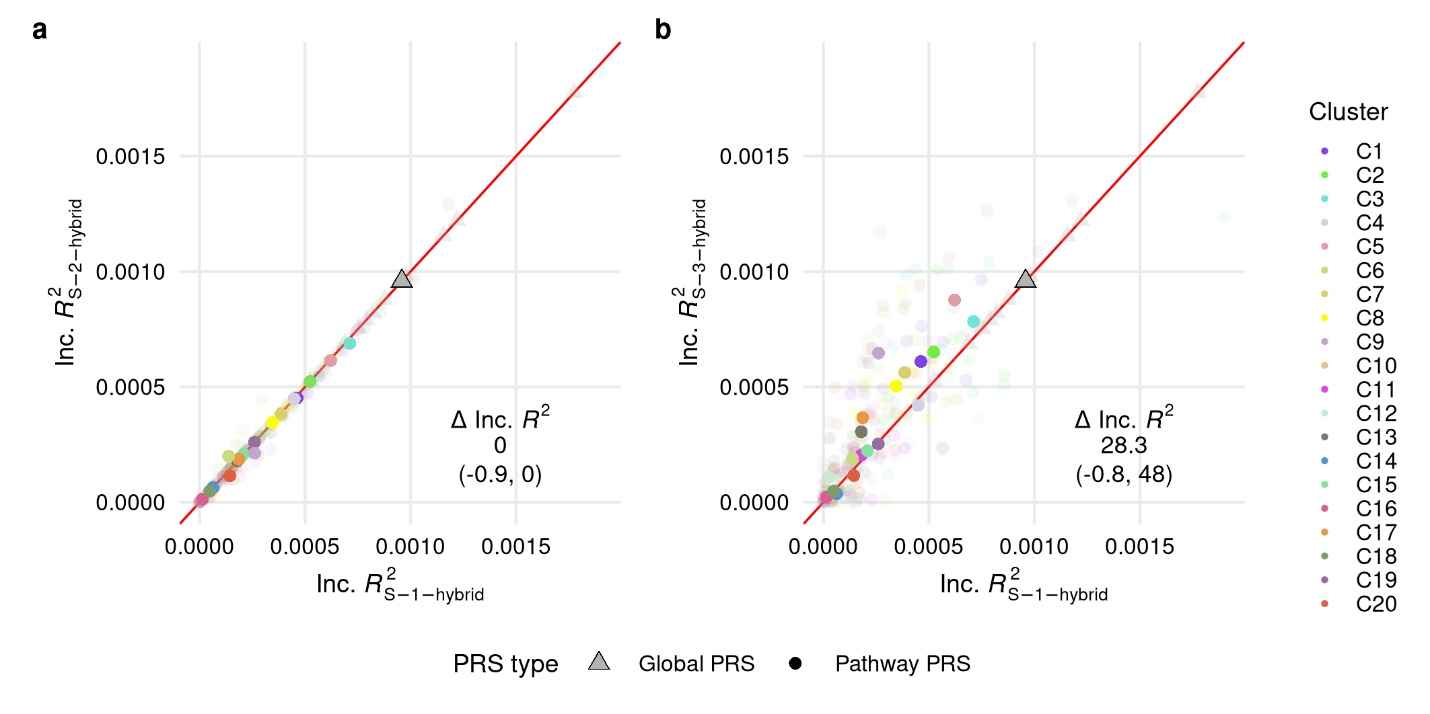

**Figure S9 |** **Hybrid C+T global and pathway-PRS predictive performance in the proxy analysis.** Performance in the UK Biobank testing sets under **(a)** S-1 vs S-2 annotation, **(b)** S-1 vs S-3 annotation.
Inc.$R_{X}^{2}$, incremental $R^{2}$ for each PRS as full model compared to covariate-only model; ∆Inc.$R^{2}$, percent change in Inc.$R^{2}$ as median (Q1, Q3); transparent points, Inc.$R^{2}$ estimates from individual holdouts; solid points, average Inc.$R^{2}$ over ten repeated holdouts.

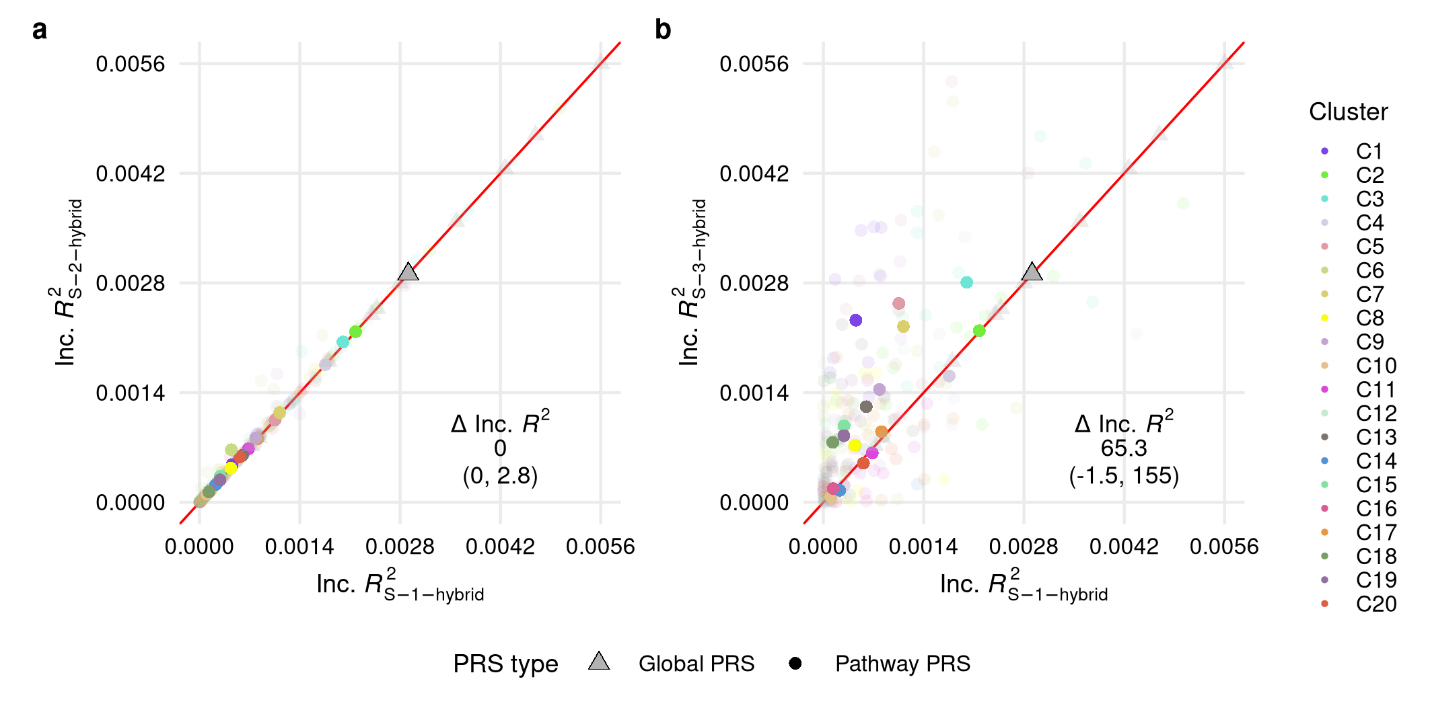

**Figure S10 |** **Hybrid C+T global and pathway-PRS predictive performance in the true case analysis.** Performance in the UK Biobank testing sets under **(a)** S-1 vs S-2 annotation, **(b)** S-1 vs S-3 annotation.

Inc.$R_{X}^{2}$, incremental $R^{2}$ for each PRS as full model compared to covariate-only model; ∆Inc.$R^{2}$, percent change in Inc.$R^{2}$ as median (Q1, Q3); transparent points, Inc.$R^{2}$ estimates from individual holdouts; solid points, average Inc.$R^{2}$ over ten repeated holdouts.

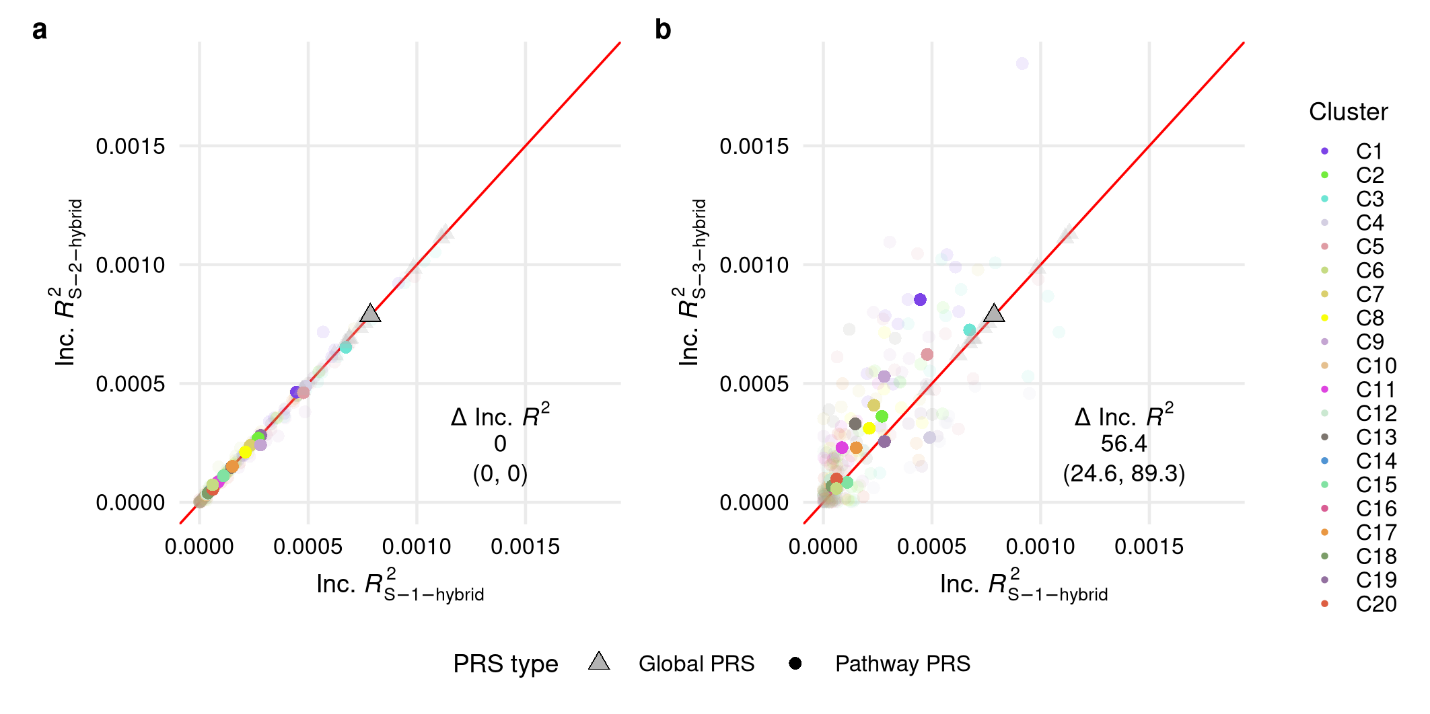

**Figure S11 |** **Hybrid C+T global and pathway-PRS predictive performance in the Wu/Marioni analysis.** Performance in the UK Biobank testing sets under **(a)** S-1 vs S-2 annotation, **(b)** S-1 vs S-3 annotation.
Inc.$R_{X}^{2}$, incremental $R^{2}$ for each PRS as full model compared to covariate-only model; ∆Inc.$R^{2}$, percent change in Inc.$R^{2}$ as median (Q1, Q3); transparent points, Inc.$R^{2}$ estimates from individual holdouts; solid points, average Inc.$R^{2}$ over ten repeated holdouts.

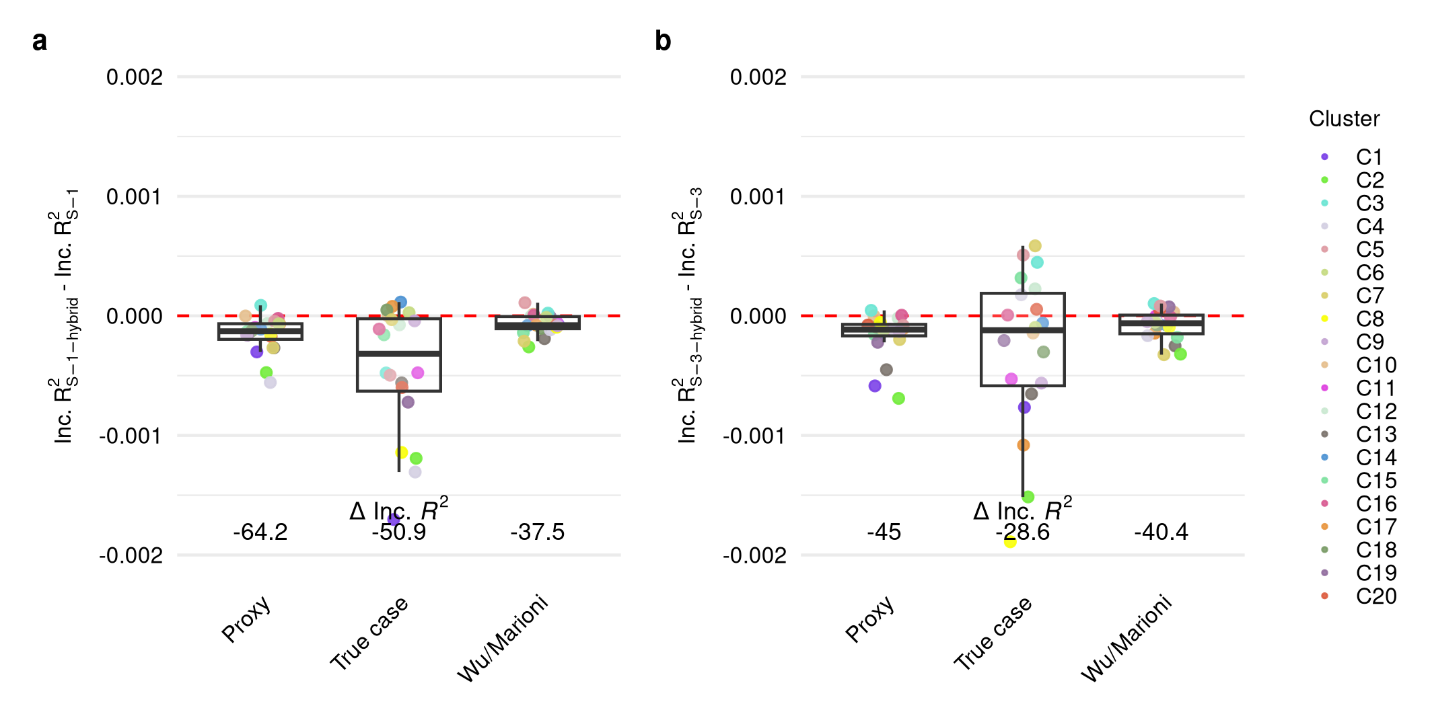

**Figure S12 |** **Hybrid pathway-PRS performance**. Difference in pathway-PRS predictive performance as averaged incremental $R^{2}$ across repeated holdout testing sets between **(a)** hybrid C+T and standard C+T based on S-1 annotation, **(b)** hybrid C+T and standard C+T based on S-3 annotation.

Training and testing conducted on (**Proxy**) proxy analysis dataset with adjustment for participant age*, sex*, number of *APOE* 𝜀4 alleles, and top PCs, (**True case**) true case analysis dataset with adjustment for participant age, sex, number of *APOE* 𝜀4 alleles, and top PCs, (**Wu/Marioni**) Wu/Marioni analysis dataset with adjustment for participant birth year, sex, maternal age, paternal age, genotyping batch, number of *APOE* 𝜀4 alleles, and top PCs.

Inc.$R_{X}^{2}$, incremental $R^{2}$ for each PRS as full model compared to covariate-only model, averaged over ten repeated holdouts; ∆Inc.$R^{2}$, median percent change in average Inc.$R^{2}$across clusters; *age and sex among proxy cases in the proxy analysis are the age and sex of the affected parent or average of affected parents.

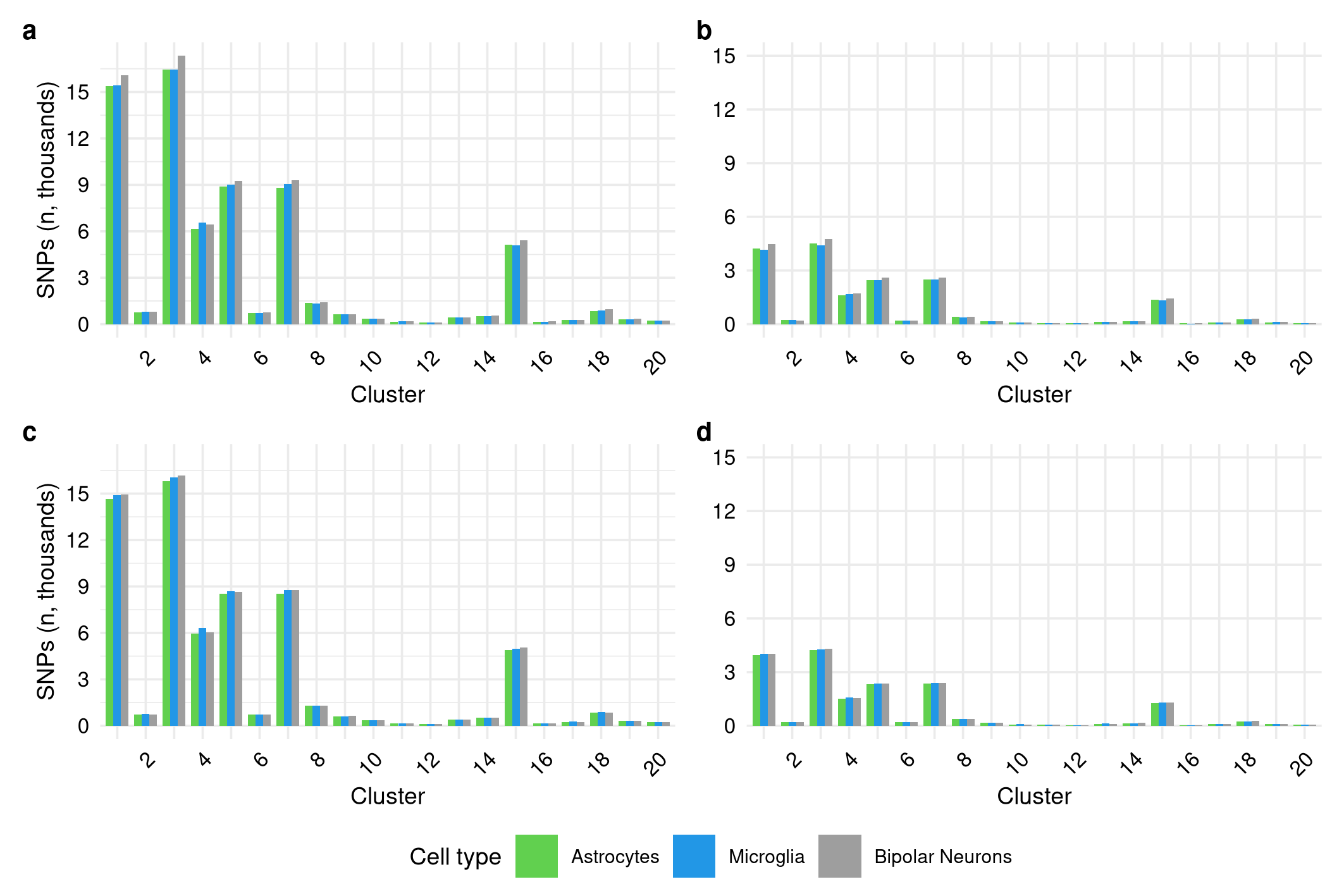

**Figure S13 | ABC-derived SNP-to-gene annotation.** Number of unique SNPs annotated to each cluster under S-3 modified with ABC-predicted enhancer-gene pairs in astrocytes, bipolar neurons, and microglia for **(a)** all predicted enhancer-gene pairs **(b)** all predicted enhancer-gene pairs with SNPs filtered to unique linkage disequilibrium (LD)-independent SNPs (*r^2^* = 0.1) **(c)** predicted enhancer-gene pairs with the max ABC Score for a given enhancer (ABC-Max) and **(d)** ABC-Max with SNPs filtered to unique linkage disequilibrium (LD)-independent SNPs (*r^2^* = 0.1).

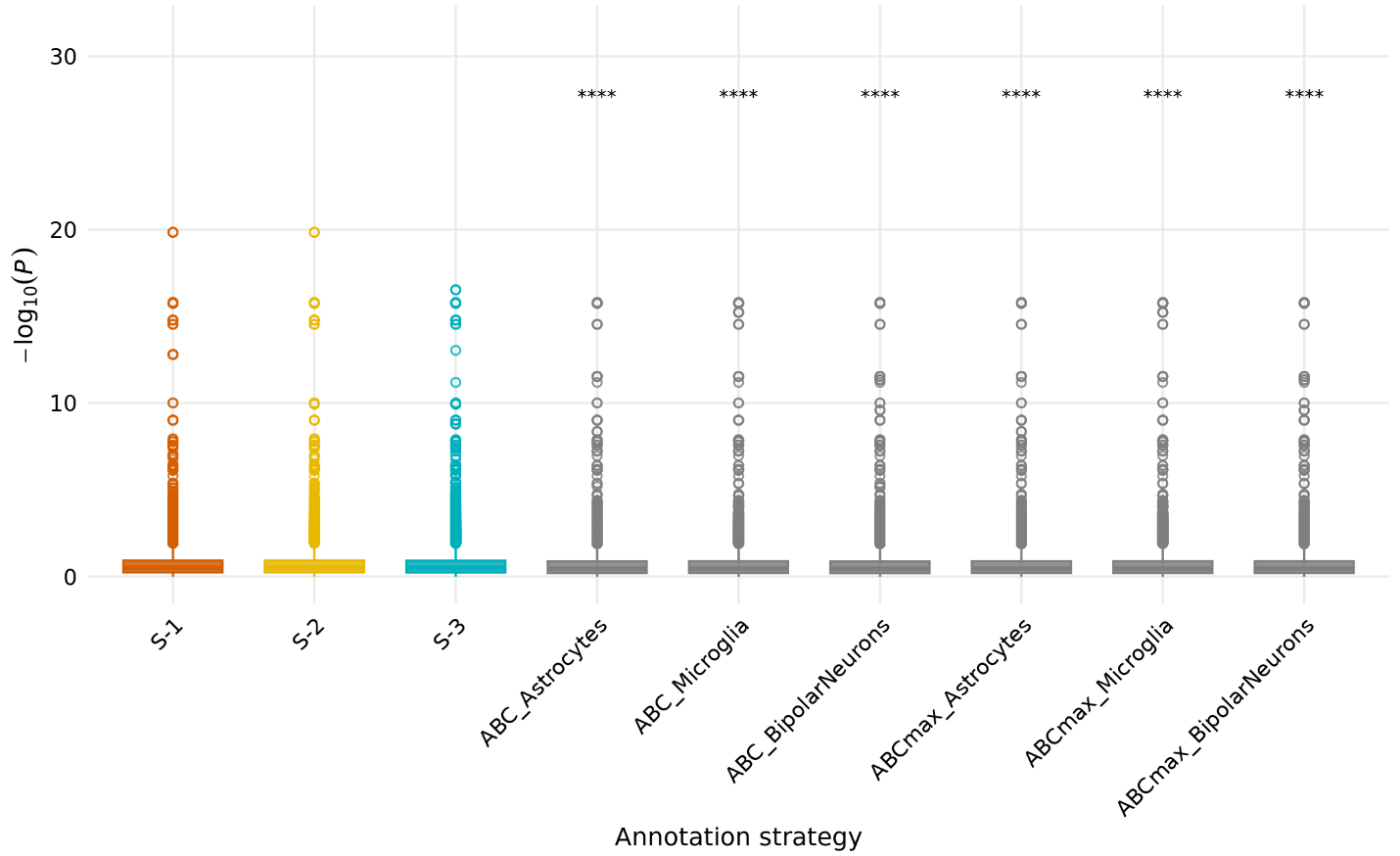

**Figure S14 |** ***P*-value distributions of LD-independent annotated SNPs across clusters by annotation strategy.** Kruskal-Wallis test *****p* ≤ 0.0001, ****p* ≤ 0.001, ***p* ≤ 0.01, **p* ≤ 0.05.

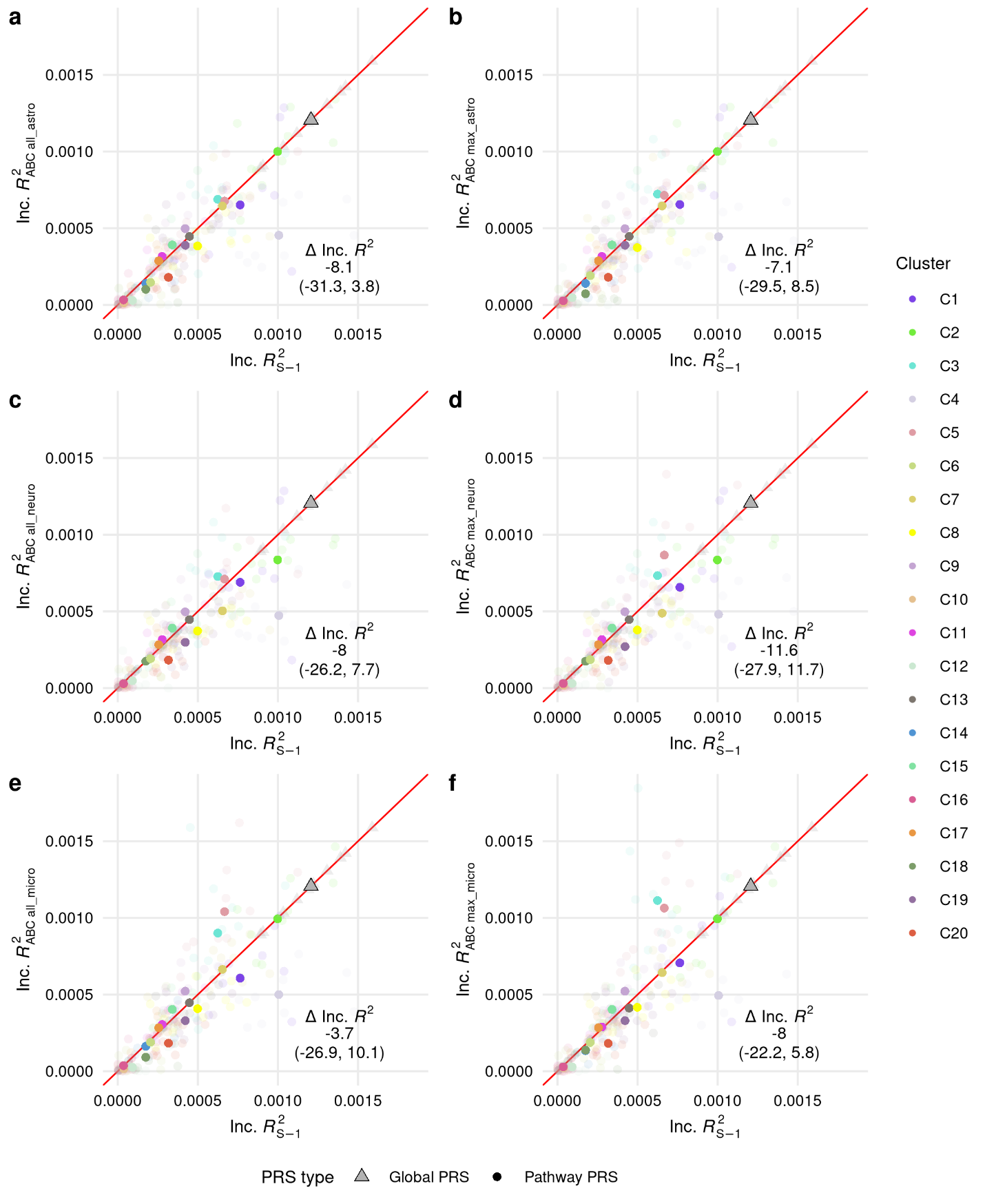

**Figure S15 | Global and pathway-PRS predictive performance in the proxy analysis.** Performance in the UK Biobank testing sets under S-1 annotation (x-axis) and modified S-3 with ABC-predicted enhancer-gene pairs (y-axis) for **(a-c)** ABC-all in astrocytes, bipolar neurons, and microglia and **(d-f)** ABC-max in astrocytes, bipolar neurons, and microglia.

Inc.$R_{X}^{2}$, incremental $R^{2}$ for each PRS as full model compared to covariate-only model; ∆Inc.$R^{2}$, percent change in Inc.$R^{2}$ as median (Q1, Q3); transparent points, Inc.$R^{2}$ estimates from individual holdouts; solid points, average Inc.$R^{2}$ over ten repeated holdouts.

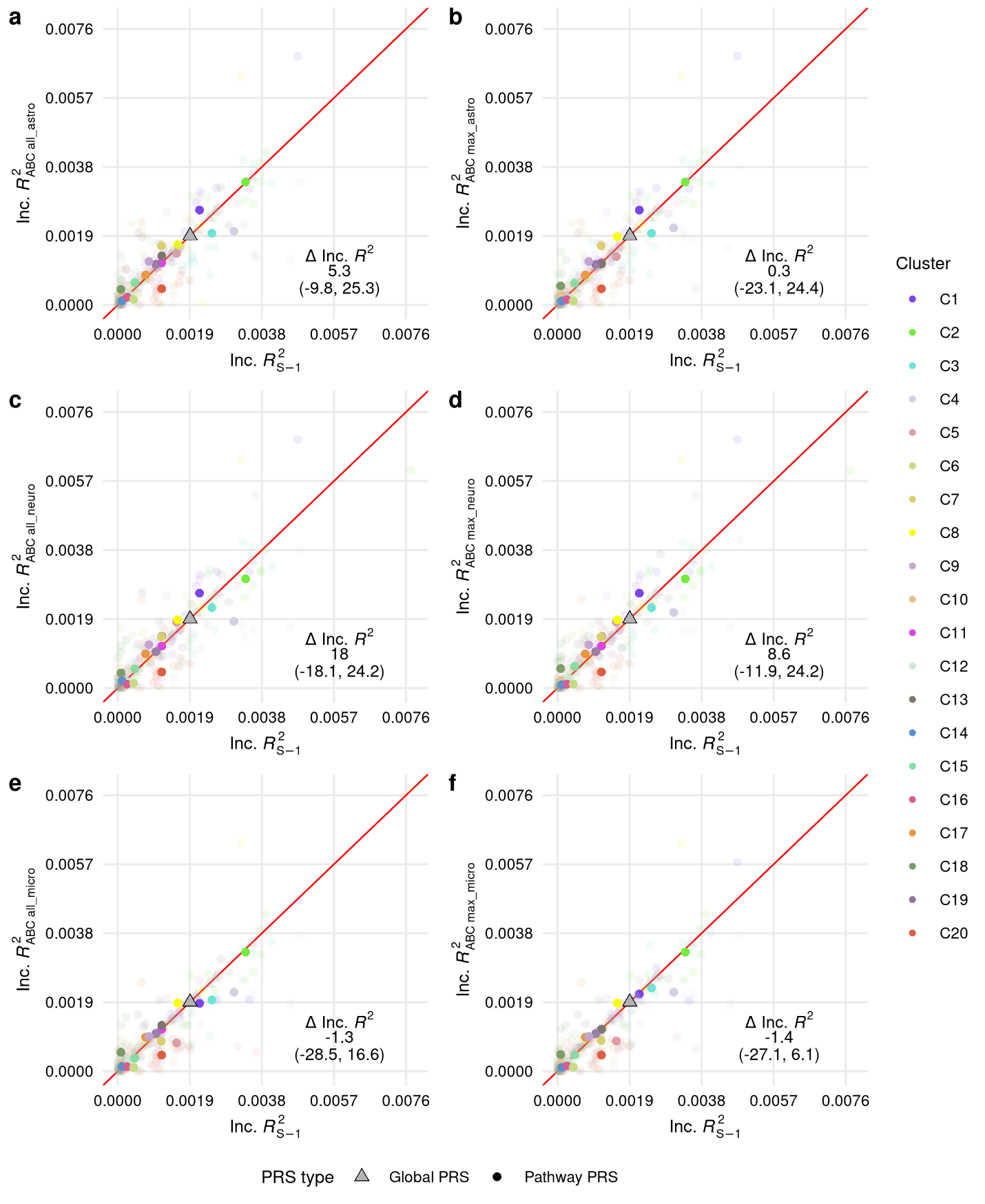

**Figure S16 | Global and pathway-PRS predictive performance in the true case analysis.** Performance in the UK Biobank testing sets under S-1 annotation (x-axis) and modified S-3 with ABC-predicted enhancer-gene pairs (y-axis) for **(a-c)** ABC-all in astrocytes, bipolar neurons, and microglia and **(d-f)** ABC-max in astrocytes, bipolar neurons, and microglia.
Inc.$R_{X}^{2}$, incremental $R^{2}$ for each PRS as full model compared to covariate-only model; ∆Inc.$R^{2}$, percent change in Inc.$R^{2}$as median (Q1, Q3); transparent points, Inc.$R^{2}$ estimates from individual holdouts; solid points, average Inc.$R^{2}$ over ten repeated holdouts.

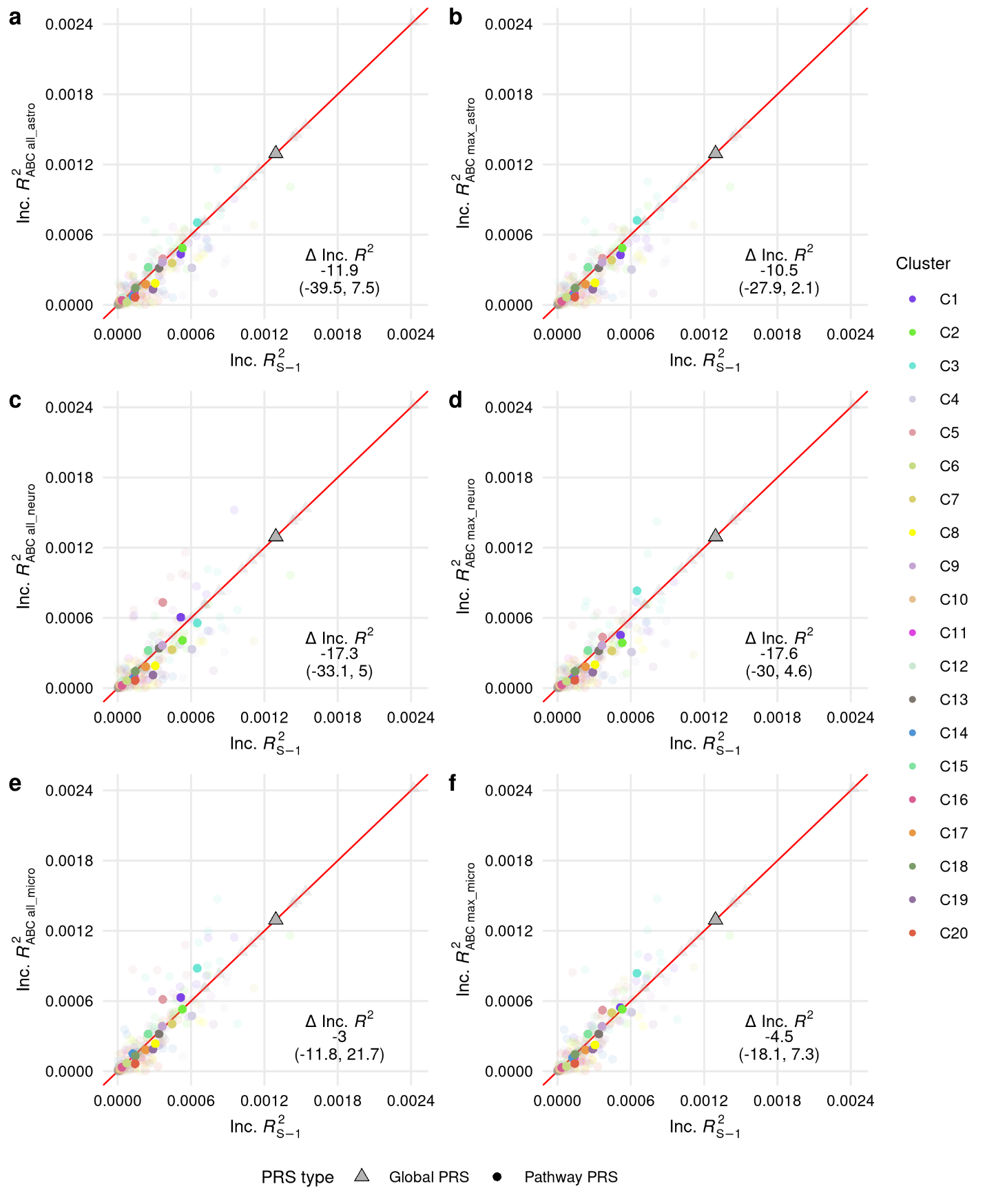

**Figure S17 | Global and pathway-PRS predictive performance in the Wu/Marioni analysis.** Performance in the UK Biobank testing sets under S-1 annotation (x-axis) and modified S-3 with ABC-predicted enhancer-gene pairs (y-axis) for **(a-c)** ABC-all in astrocytes, bipolar neurons, and microglia and **(d-f)** ABC-max in astrocytes, bipolar neurons, and microglia.
Inc.$R_{X}^{2}$, incremental $R^{2}$ for each PRS as full model compared to covariate-only model; ∆Inc.$R^{2}$, percent change in Inc.$R^{2}$as median (Q1, Q3); transparent points, Inc.$R^{2}$ estimates from individual holdouts; solid points, average Inc.$R^{2}$ over ten repeated holdouts.

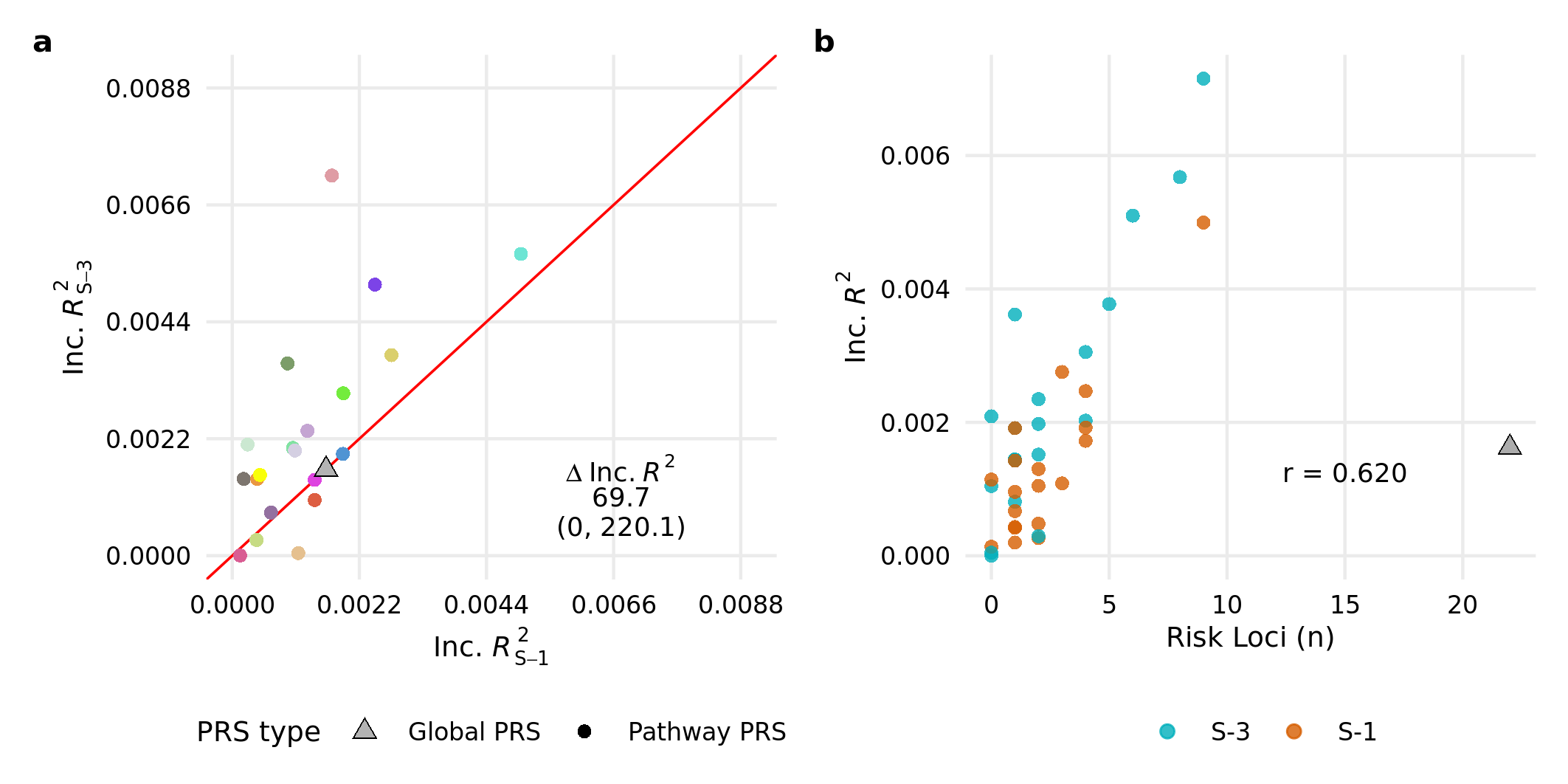

**Figure S18 |** **Global and pathway-PRS trained on the UK Biobank true case analysis dataset and calculated in the ADGC. (a)** predictive performance across S-1 and S-3 annotation, and **(b)** number of genome-wide significant AD risk loci represented in the PRS model compared to predictive performance.
Inc.$R_{X}^{2}$, incremental $R^{2}$ for each PRS as full model compared to covariate-only model; ∆Inc.$R^{2}$, percent change in Inc.$R^{2}$ as median (Q1, Q3); r, spearman correlation between number of risk loci and incremental $R^{2}$; Genome-wide significant risk loci obtained from Kunkle et al 2019 Table S8.

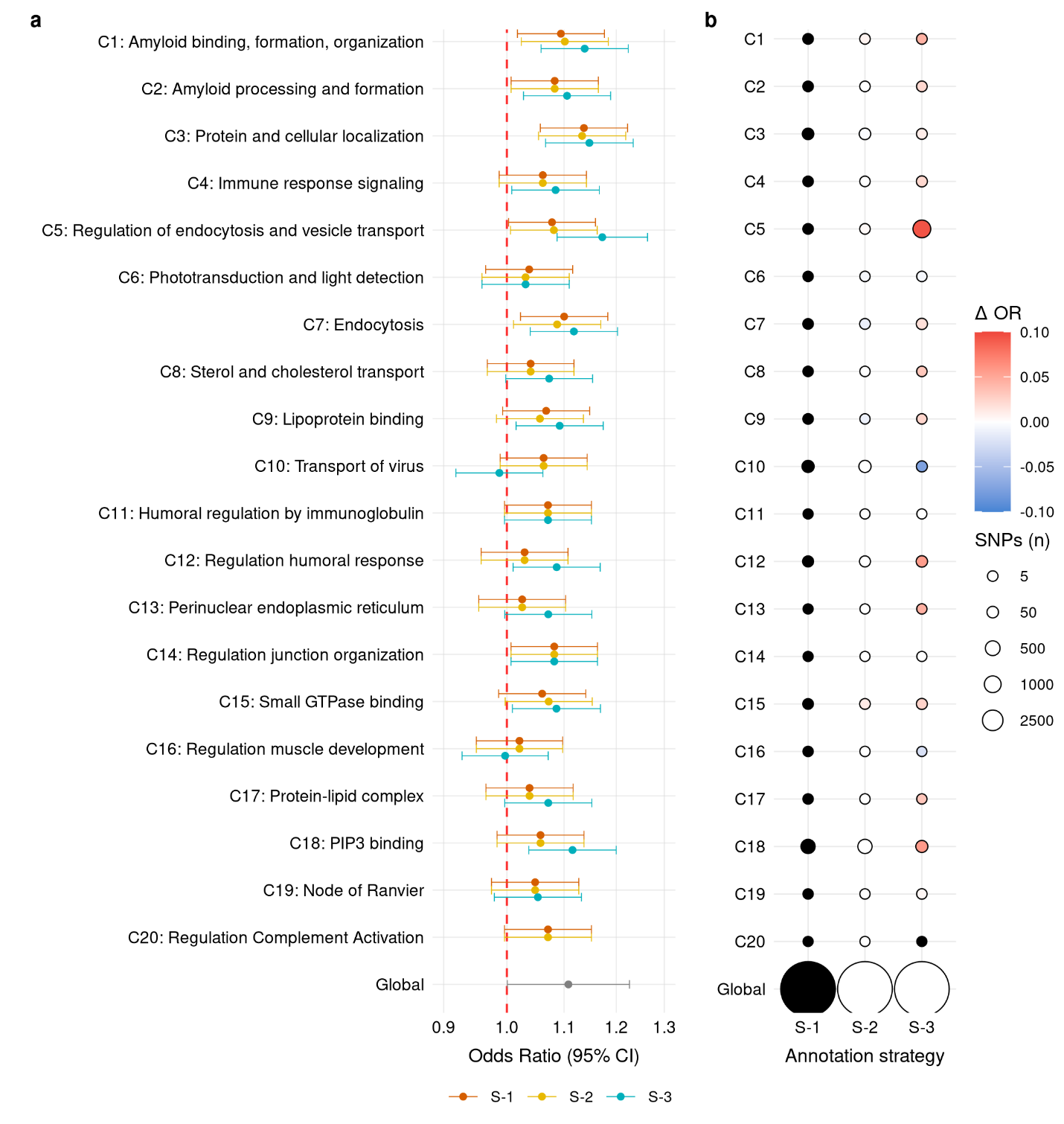

**Figure S19 |** **Global and pathway-PRS trained on the UK Biobank true case analysis dataset and calculated in the ADGC. (a)** association with AD status across annotation strategies, and **(b)** relative size of PRS models as the number of included SNPs and relative strength of association with AD status across annotation strategies.
∆OR vs S-1, difference in estimated odds ratio for the association with AD status from the S-1 pathway-PRS odds ratio.

C20 S-3 pathway-PRS removed from (a) due to extreme confidence intervals, OR: 2.29 95% CI: 0.82, 6.38.
